# Defining Key Deprescribing Measures from Electronic Health Data: A Multisite Data Harmonization Project

**DOI:** 10.1101/2023.11.06.23298060

**Authors:** Sascha Dublin, Ladia Albertson-Junkans, Thanh Phuong Pham Nguyen, Juliessa M. Pavon, Susan N. Hastings, Matthew L. Maciejewski, Allison Willis, Lindsay Zepel, Sean Hennessy, Kathleen B. Albers, Danielle Mowery, Amy G. Clark, Sunil Thomas, Michael A. Steinman, Cynthia M. Boyd, Elizabeth A. Bayliss

**Affiliations:** Kaiser Permanente Washington Health Research Institute, Seattle, WA; University of Washington Department of Epidemiology, Seattle, WA; Kaiser Permanente Bernard J. Tyson School of Medicine, Pasadena, CA; University of Pennsylvania Perelman School of Medicine, Philadelphia, PA; Duke University School of Medicine, Durham, NC; Durham VA Healthcare System, Geriatrics Education Research and Clinical Centers; Durham VA Center of Innovation to Accelerate Discovery and Practice Transformation; Duke University, Department of Population Health Sciences; Kaiser Permanente Colorado Institute for Health Research, Aurora, CO; Division of Geriatrics, University of California San Francisco, San Francisco, CO; San Francisco Veterans Administration Medical Center, San Francisco, CA; Division of Geriatric Medicine and Gerontology and Center for Transformative Geriatric Research, Johns Hopkins University School of Medicine, Baltimore, MD; Department of Family Medicine, University of Colorado School of Medicine, Aurora, CO

**Author notes:** Corresponding author: Dr. Sascha Dublin, 1730 Minor Avenue, Suite 1600, Seattle, WA, 98101. Disclosures (all funding sources): This work was funded by National Institute on Aging grants R24AG064025 and 1K23AG058788-03 (Pavon) and US Department of Veterans Affairs grant RCS 10-391 (Maciejewski). The contents do not represent the views of the U.S. Department of Veterans Affairs or the United States Government. Prior presentations: Portions of this paper were presented at the US Deprescribing Research Network Annual Meeting in Long Beach, California in May 2023; at the International Conference on Pharmacoepidemiology in Halifax, Nova Scotia in August 2023; and at the National Association of Primary Care Research Group annual meeting in San Francisco, California in November 2023.

**Keywords:** deprescribing, measurement, electronic health records, benzodiazepines, medication discontinuation

## Abstract

**Background:** Deprescribing, or systematically stopping or reducing risky or unneeded medications, could improve older adults’ health. Electronic health records (EHR) hold promise for supporting deprescribing studies, but there are currently no standardized measures for key variables. With benzodiazepines and other sedative-hypnotics (Z-drugs) as a case study, we developed and examined EHR-based definitions for chronic medication use and discontinuation.

**Methods:** We conducted a retrospective cohort study set within 5 U.S. healthcare systems. The study population was adults age 65+ from 2017-2019 with chronic benzodiazepine or Z-drug use, without dementia or serious mental illness. We used EHR data for medication orders and dispensings to define key variables, including chronic benzodiazepine/Z-drug use and discontinuation. We explored definitions for discontinuation based on 1) gaps in medication availability during follow-up (no active order/dispensing) or 2) not having medication available at a fixed time point. We examined the impact of varying gap length from 30 to 180 days, accounting for stockpiling, and requiring a 30-day period without orders/dispensings (“halo”) around the fixed time point. We also compared results from medication orders vs. dispensings for the same population.

**Results:** 1.6-2.6% of older adults had chronic use of a benzodiazepine or Z-drug. Depending on the definition and site, the proportion discontinuing use over 12 months ranged from 6% to 49%. Requiring a longer gap in orders/dispensings or a 30-day “halo” around a fixed time point resulted in lower estimates. Orders data were less likely to identify discontinuation than dispensing data.

**Conclusions:** Requiring a medication gap of ≥90 days or a 30-day period with no orders/dispensings around a fixed time point may improve the likelihood that an outcome represents true discontinuation. Orders data appear to underestimate discontinuation compared to dispensing data. More work is needed to adapt and test the proposed definitions for other drug classes and care settings.

**Impact Statement:** We certify that this work is novel. Prior papers have identified a need for greater standardization of definitions for medication exposure and discontinuation in deprescribing studies. To our knowledge, no prior paper has systematically examined the construction of deprescribing variables from electronic health data. This is the first paper to present standardized definitions for variables needed for deprescribing studies based on electronic health data, to implement these definitions in multiple healthcare systems and data types, and to examine their performance. This is also the first paper to examine the impact of using medication orders vs. dispensing data to define key deprescribing variables.

**Key points:**

- Using benzodiazepines and Z-drugs as a case study, we developed and implemented standardized definitions for key variables needed for deprescribing studies using electronic health records data from 5 diverse U.S. healthcare systems.
- Requiring a gap of ≥90 days without an active order/dispensing or, for a fixed time point, requiring a period of ≥30 days surrounding it with no order/dispensing will likely increase the accuracy of identifying true medication discontinuation.
- Applying definitions to medication orders data generated higher estimates of chronic use and lower estimates of medication discontinuation than dispensing data.

**Why does this paper matter?:** The use of standardized variable definitions in deprescribing studies will improve the ability to synthesize data and compare results between studies, advancing knowledge and supporting more evidence-based guidelines for clinical care.

## Introduction

Polypharmacy is common in older adults and has been associated with many adverse outcomes.[1–4] A potential solution is deprescribing – systematically stopping or tapering medications that are risky or no longer needed, with ongoing monitoring.[5, 6] There are important gaps in the evidence about deprescribing, including its impact on health outcomes.

Electronic health records (EHR) data can support evidence generation, for instance by supporting efficient recruitment and outcome ascertainment for randomized controlled trials (RCTs). They can also be used for observational studies. Benefits include access to large and more representative populations.

Deprescribing studies have used heterogeneous definitions for key outcome variables, including chronic medication use and discontinuation, hindering comparison and synthesis of results.[7] EHR-based definitions for these variables have not been systematically proposed or examined. To address this gap, the US Deprescribing Research Network convened a workgroup to develop and implement standardized EHR-based definitions for key variables for deprescribing studies.

## Methods

### Overview

Researchers from 5 U.S. healthcare systems met regularly, selecting high-priority variables and collaboratively developing definitions that each site operationalized. We carried out a retrospective cohort study, identifying individuals with chronic medication use and following them to identify discontinuation. As a case study, we chose benzodiazepines and related sedative-hypnotics (zolpidem, eszopiclone, and zaleplon, or “Z-drugs”), common targets for deprescribing. To provide a framework, we emulated some elements of D-PRESCRIBE, a community-based deprescribing RCT.[8]

### Study settings and data sources

Participating healthcare systems (Appendix Table 1) included 2 integrated healthcare systems, regions of Kaiser Permanente (KP); two academic healthcare systems, the University of Pennsylvania and Duke University; and the Durham Veterans Affairs Health Care System (VAHCS). The first 4 use Epic EHRs. All systems have EHR data for demographic characteristics, visits, diagnoses, and medication orders. The KP regions also have claims data, including health plan enrollment and pharmacy dispensings. Medication orders come from the EHR and show prescriptions ordered by a clinician, while dispensing data are generated by pharmacies as part of filling these prescriptions. One academic site obtained dispensing data from a vendor, Surescripts, while the other had only orders data.

### Study population

Inclusion and exclusion criteria were modeled after D-PRESCRIBE (see Appendix Table 2). We included people aged 65 and older who engaged with our healthcare systems from 2017- 2019. We required chronic use of a benzodiazepine or Z-drug in 2018 (definitions follow). We excluded people with serious mental illness or significant cognitive impairment and those needing an interpreter.

### Defining chronic benzodiazepine or Z-drug use

Table 1 provides definitions for key variables. We also provide a Programmers’ Guide (Supplemental Appendix Text) with practical and technical guidance for implementing the definitions. Sites with dispensing data defined chronic benzodiazepine/Z-drug use as having ≥3 dispensings and ≥45 days’ supply within 100 days. The rationale for the days’ supply requirement is that a more stringent definition of chronic use will identify individuals at higher risk of harm and make it easier to ascertain discontinuation accurately. When using orders data to define chronic use, we required ≥3 orders within 100 days, counting the initial order and any refills included. Implicitly, these definitions assume that a typical dispensing/prescription is for ≤30 days’ supply. The orders-based definition does not include a days’ supply requirement because orders generally lack this element. When defining chronic use in 2018, we considered dispensings/orders from 2017 and 2018.

**Table 1:**
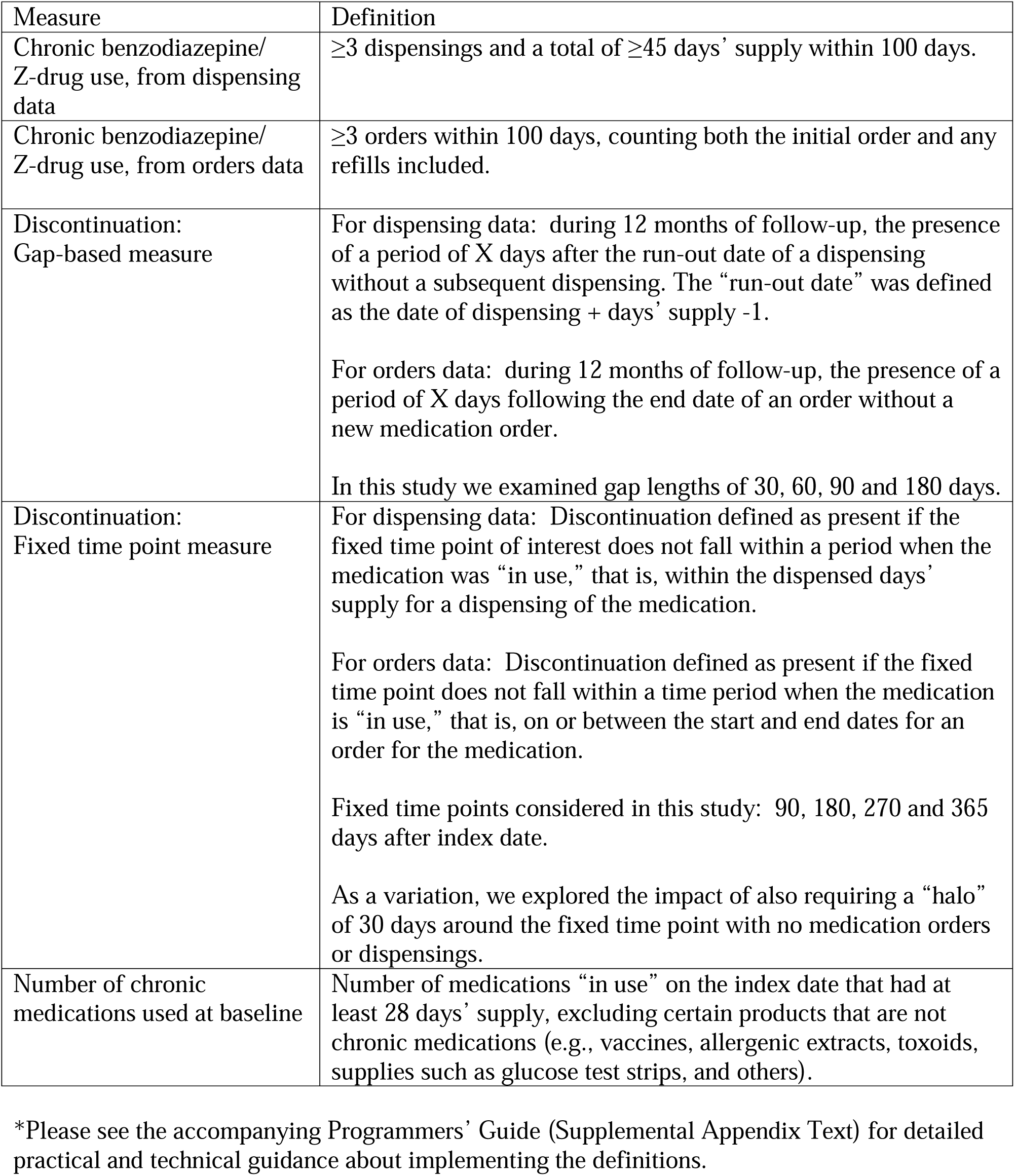
Definitions for Deprescribing Measures*.

Individuals were assigned an “index date” which separated the baseline period, during which chronic use was assessed, from the follow-up period, during which discontinuation was assessed. In dispensing data, the index date was the date of the dispensing that qualified a person as having chronic use (i.e., the third dispensing or the dispensing that led to ≥45 total days’ supply). It is challenging to assign an index date from orders data because people may qualify as having chronic use based on allowed refills, which do not have a set start date. Thus, for people meeting chronic use criteria in 2018 based on orders, we assigned 1/1/2019 as the index date.

### Baseline characteristics

We extracted data about demographics, diagnoses, and medication use (Appendix Table 3). We ascertained opioid use because these medications are often used with benzodiazepines, and concomitant use increases the risk of adverse outcomes. Chronic opioid use was identified using the same definition as for benzodiazepines/Z-drugs.

We measured the number of chronic medications other than benzodiazepines/Z-drugs on the index date. First, we identified those where the index date fell within the medication’s days’ supply (using dispensing data) or on or between an order’s start and end date. For dispensings, we included only those with ≥28 days’ supply. We excluded products that are not chronic medications (e.g., vaccines). We then counted the number of distinct national drug codes for dispensing data or generic medication names (for orders).

### Defining medication discontinuation

We took two conceptual approaches (Table 1): 1) determining whether a person had stopped use for a set *duration of time* at any point during a specified follow-up period (“gap-based” definition) and 2) determining whether the person had stopped using the medication at a *fixed time point* during follow-up. We followed individuals for 12 months after the index date.

For dispensings, a medication was considered “in use” from the dispensing date through the run-out date, defined as dispensing date + days’ supply – 1 (assuming the first dose is taken on the dispensing date). If there was not another dispensing within 30 days after the run-out date, we considered this a 30-day gap. Similarly, we identified gaps of 60, 90, or 180 days. As a variation, we defined gaps accounting for medication stockpiling.

For orders, a medication was considered “in use” from the order start date through the end date. The order end date accounts for allowed refills (essentially, assuming all allowed refills were utilized). It can be retroactively modified by various events, such as the patient reporting that they stopped the medication.

For the fixed time point definition, we assessed whether a person appeared to have the medication on hand at a given time point (Appendix Figure 1a). If the time point did not fall within a period when the medication was in use, this was considered discontinuation. We defined discontinuation at days 90, 180, 270 and 365 of follow-up. We realized that for a person continuing use, a fixed time point could fall within a brief gap between two dispensings. Thus, as a variation, we required that the fixed time point fall within a 30-day period with no dispensings or orders (“halo”; see Appendix Figure 1b).

We created variables for discontinuation using the gap-based and fixed time point approaches and compared the results.

### Comparing results from medication orders versus dispensings

Orders and dispensing data differ in important ways that could affect measurement.

Orders lack information about days’ supply, and it is impossible to determine whether allowed refills were dispensed or when. The order end date is calculated assuming all refills were used, potentially overestimating use. We utilized data from KPWA, which has both orders and dispensing data, to define chronic benzodiazepine/Z-drug use and discontinuation using both data types for a set population and examined agreement.

## Results

In 2018, the prevalence of chronic use of benzodiazepines/Z-drugs ranged from 1.5% to 2.6% across sites (Appendix Figure 2). Appendix Figures 3a-3f show the impact of inclusion and exclusion criteria. People with chronic benzodiazepine/Z-drug use had a mean age of 71.5-73.3 years, about two-thirds were female (except at the VAHCS), and most were white (Appendix Table 3). Between 31-66% had a diagnosis of anxiety in the EHR and 4-31% a diagnosis of insomnia.

Across sites, the proportion of people with a gap in use of ≥30 days during follow-up varied from 13-49% and for gaps ≥180 days, from 6-15% (Figure 1). As the gap length increased, the proportion discontinuing decreased. Site 5, which used orders data, had the lowest apparent discontinuation rates.

**Figure 1.**
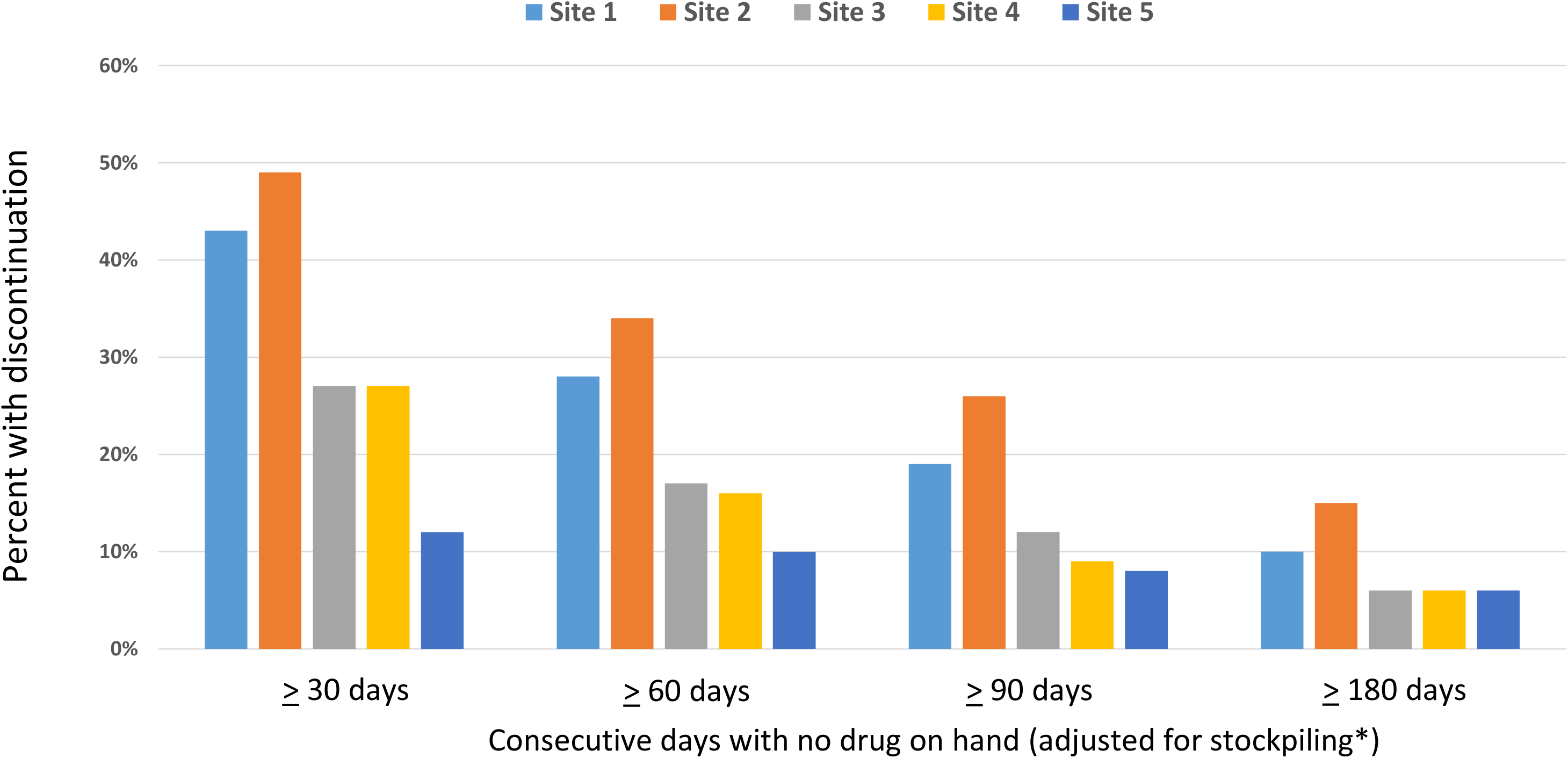
Prevalence of medication discontinuation during 12 months of follow-up across sites, using a gap-based definition and varying gap length.

With the fixed time point definition, the proportion of individuals who discontinued benzodiazepine/Z-drug use ranged from 9-32% at 180 days after the index date and from 20-36% at 365 days (Figure 2). Requiring a 30-day “halo” with no order/dispensing around the fixed point led to substantially lower estimates. Discontinuation rates were slightly lower after adjustment for stockpiling (Appendix Figure 4), but for longer gaps, this adjustment mattered less.

**Figure 2.**
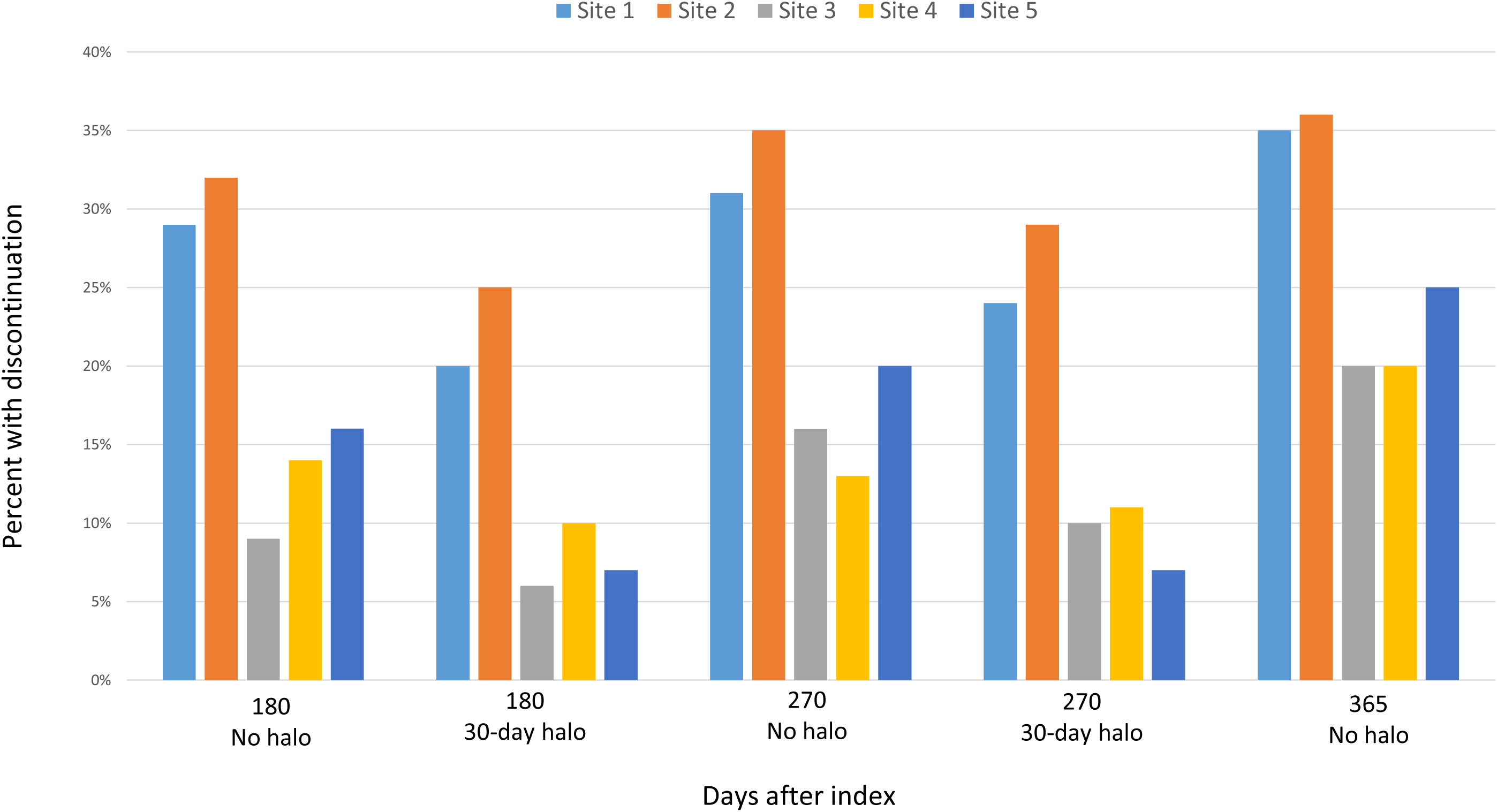
Prevalence of medication discontinuation during 12 months of follow-up across sites, using the fixed time point definition. Estimates for Sites A-D are adjusted for stockpiling. Site E estimates are based on orders data and given challenges in measuring order end dates, there was no adjustment for stockpiling.

In an analysis of individuals at one site with both orders and dispensing data, far more people appeared to have chronic benzodiazepine/Z-drug use based on orders than dispensings (Appendix Table 4a). Of those with chronic use defined from orders, only 56% also qualified based on dispensings. The discrepancy was greatest among individuals who qualified based on a single order (Appendix Table 4b). Discontinuation rates derived from dispensings were substantially higher than from orders (Appendix Table 5); for example, at 180 days after index date, the discontinuation rate was 32% based on dispensings vs. 20% based on orders.

## Discussion

We found that at 5 U.S. healthcare systems in 2018, 1.5-2.6% of older adults were using benzodiazepines or Z-drugs chronically. For comparison, a U.S. study using dispensing data found that in 2008, 2.7% of people aged 65-80 had long-term benzodiazepine use,[9] while in Canada in 2006, 3.5% of older adults had long-term use.[10]

This multisite investigation yielded three key insights. First, discontinuation rates over 12 months of follow-up ranged from 6 to 49% depending on the definition; requiring a longer gap to qualify as discontinuation resulted in a lower discontinuation rate. Our results suggest that gaps of 30 or 60 days may be less likely to represent true discontinuation than longer gaps. Second, when defining discontinuation at a fixed time point, requiring a 30-day “halo” period resulted in lower discontinuation rates. This indicates that many people received subsequent orders/fills shortly after the time point, suggesting they did not truly discontinue. Third, data from orders classified more people as having chronic use and identified fewer discontinuations than dispensing data. Using orders data for this purpose poses challenges, including ambiguity about refill usage. Orders may remain active long after use has stopped. Thus, dispensing data are likely more precise for these purposes.

This is the first paper to examine the construction of deprescribing variables from EHR data systematically. Our results align with some prior findings. Niznik et al. examined bisphosphonate discontinuation and recommended requiring longer gaps (≥90 days).[11] Another study compared definitions of opioid use from orders vs. dispensing data. They had orders data from a single healthcare system and found that dispensing data yielded higher opioid use estimates than orders (the reverse of our findings).[12] These results illustrate the challenges of using orders data, in this case the potential to underestimate use due to fragmentation of care.

Strengths of this project include the involvement of 5 healthcare systems representing different U.S. regions, patient populations, and types of data. We examined several definitions for medication discontinuation and developed a programmers’ guide (see Appendix).

Benzodiazepines and Z-drugs are controlled substances and are the subject of many policies, guidelines and regulations. Some of these limit prescriptions to 30 days’ supply[13], and at our sites most dispensings were for ≤30 days. This influenced our algorithms. Other chronic medications are often prescribed with 90 days’ supply, which may affect the transferability of our definitions. Also, benzodiazepines/Z-drugs are often used sporadically, making it harder to identify discontinuation compared to daily medications. We lacked a gold standard against which we could evaluate our definitions’ performance. Our study did not investigate the reasons for discontinuation, so we could not distinguish between clinically recommended discontinuation and patient-driven discontinuation without clinical support.

Additional research is needed to understand the intent behind observed discontinuations and to develop EHR-based definitions for dose reduction or tapering. Our proposed definitions should be adapted and tested for other chronic medications and care settings.

In conclusion, it is feasible to develop and apply standardized, EHR-based definitions for deprescribing variables in diverse settings. When selecting variable definitions, researchers should consider the potential impacts of misclassification given their study design and aims. Our goal is to provide guidance for these decisions and to support greater standardization of measures, improving the evidence base for deprescribing.

## Supporting information

Supplemental Text Programmers' Guide

## Data Availability

Data produced in the present study are not available for sharing because they come from electronic health records governed by United States HIPAA regulations and participants did not provide consent for data sharing (data were obtained with a waiver of consent.)

## Acknowledgments

Conflicts of interest: SD has received funding from GSK. SH has consulted for or received funding from Pfizer, Johnson & Johnson, Novo Nordisk, Arbor Pharmaceuticals, the Medullary Thyroid Cancer Consortium (Novo Nordisk, AstraZeneca, GlaxoSmithKline and Eli Lilly), Biogen, Intercept Pharmaceuticals, Provention Bio, bluebird bio, and Amylyx Pharmaceuticals. TPPN was a member of the 2022-2023 Junior Investigator Intensive Program of the US Deprescribing Research Network, which is funded by the National Institute on Aging, and has received support from Acadia Pharmaceuticals. MM owns stock in Amgen. Other authors have no conflicts to report.

## Author Contributions

Study concept and design: SD, LAJ, TPPN, JMP, SNH, MLM, AGC, SH, AW, KBA, DM, EAB Acquisition of data: LAJ, KBA, DM, LZ, ST

Analysis and interpretation of data: all authors Preparation of manuscript: all authors

Sponsor’s Role: The sponsor did not play a role in study design, methods, data collection, analysis, or preparation of the manuscript.

## List of Supplemental Material for Online Publication

Supplemental Appendix Text: Programmers’ Guide. Provides detailed technical and practical guidance for working with orders and dispensing data to implement the definitions of deprescribing variables presented in this paper.

Appendix Table 1: Descriptions of healthcare systems participating in Data Harmonization Workgroup

Appendix Table 2: Definitions of inclusion and exclusion criteria and lists of diagnosis codes and medications

Appendix Table 3: Characteristics of people aged 65 or older with chronic benzodiazepine or Z-drug use at 5 US healthcare systems

Appendix Table 4a: Comparing chronic use defined from medication orders vs. dispensings for the same population

Appendix Table 4b: Comparing chronic use defined from medication orders vs. dispensings, stratified by number of medication orders

Appendix Table 5: Comparing measures of discontinuation defined from orders vs. dispensing data for the same population

Appendix Figure 1a: Illustration of fixed time point definition of discontinuation, without halo

Appendix Figure 1b: Illustration of fixed time point definition of discontinuation, with halo. In the top row, representing no discontinuation, there are dispensings or orders within the 30-day halo period around the fixed point. In the bottom line, representing discontinuation, there are no dispensings or orders within the 30-day halo period around the fixed point. Note: the 30-day period does not have to be symmetric around the fixed point; it represents any 30-day period that contains the fixed point within it.

Appendix Figure 2. Prevalence of Benzodiazepine and Z-drug Use in People Aged ≥65 Within 5 U.S. Healthcare Systems, 2018.

Appendix Figures 3a-3f: Results of Applying Inclusion and Exclusion Criteria at Each Site Appendix Figure 4: Impact of Adjusting for Medication Stockpiling on Estimated Discontinuation Rates (Using Gap-Based Definition)

