## Supplemental Text Programmers' Guide for "Defining Key Deprescribing Measures from Electronic Health Data: A Multisite Data Harmonization Project"

**Supplemental Appendix Text: Programmers Guide to Harmonizing Multisite Data
for Medication Deprescribing Research**

This Guide is for programmers and data specialists who extract and prepare medication data from clinical and administrative data sources. It may also be useful for researchers and their clinical collaborators in preparing grant applications, choosing study designs, or developing quality improvement measures for clinical operations and data systems.

The Guide is organized into six steps:

1. **Get to know your landscape**: An overview of the data domains (e.g., medication, enrollment, utilization, etc) often used in deprescribing research with a focused look at medication data.
2. **Get to know your data**: Questions to guide data exploration and documentation for cross-site understanding.
3. **Choose your data**: What to consider when deciding which data to use and why.
4. **Pull your data**: What to consider when pulling local medication data and how.
5. **Harmonize your data**: Transforming local medication data into common data elements.
6. **Build your measures**: Operationalizing concepts from common data elements.

These steps are presented sequentially but are interdependent. Data harmonization is an iterative and collaborative process that requires reconsidering each step as your team learns from the other steps. Iteration and collaboration do not have a dedicated section in this guide but should be infused throughout every step.

**Step 1: Get to know your landscape**

An essential step in any multi-site study is understanding the data available at each site. We used a data matrix like the one below (Table 1) to document this cross-site discovery work. This is a great opportunity to engage data partners and kick-start collaboration. Data partners should be encouraged to contribute to the development of this table as well as its use.

*Table 1.* Data Matrix

| **Legend**  x = raw data, well-populated  o = raw data, inconsistently populated  d = not raw but can be derived  na = not available | | **Site** | | | | |
| --- | --- | --- | --- | --- | --- | --- |
|  |  | **A** | **B** | **C** | **D** | **E** |
| Medication Data | |  |  |  |  |  |
|  | Medication orders | x | X | na | na | X |
|  | Pharmacy dispensings | x | X | x | X | na |
|  | NDC code | x | X | x | na | na |
|  | GPI code | x | X | na | na | na |
|  | Generic name | x | X | x | X | X |
|  | Administration route | x | X | x | na | na |
|  | Dosage form | x | X | x | na | na |
|  | Quantity | x | X | x | na | na |
|  | Medication start date | x | X | x | X | X |
|  | Medication end date | d | D | d | na | X |
|  | Duration | x | x | x | na | D |
| Enrollment Data | |  |  |  |  |  |
|  | Enrollment start date | x | x | na | X | na |
|  | Enrollment end date | x | x | na | X | na |
|  | Insurance type | x | x | na | x | na |
| Encounter Data | |  |  |  |  |  |
|  | Date of encounter | x | x | x | x | X |
|  | Encounter setting | x | x | x | x | X |
| Diagnosis Data | |  |  |  |  |  |
|  | Date of diagnosis | x | x | x | x | X |
|  | Diagnosis code | x | x | x | x | X |
|  | Diagnosis position | x | x | x | x | X |
| Demographic Data | |  |  |  |  |  |
|  | Date of birth | x | x | x | x | X |
|  | Sex | x | x | x | x | X |
|  | Gender | o | o | na | na | na |
|  | Primary language | x | x | x | x | X |
|  | Race | x | x | x | x | X |
|  | Ethnicity | x | x | na | na | na |
| Vital Signs Data | |  |  |  |  |  |
|  | BMI | x | x | x | x | X |
|  | Smoking status | x | x | na | na | na |
| Death Data | |  |  |  |  |  |
|  | Date of death | x | x | na | na | na |
| Site-specific exclusions | |  |  |  |  |  |
|  | Does not want to participate in research | x | x | na | na | na |
|  | Geographic location precludes research participation | | | |  | X |

Abbreviations: NDC, National Drug Code; GPI, Generic Product Identifier; BMI, body mass index

We included the full matrix to illustrate the variety of data domains that might be used in deprescribing studies. However, this guide focuses on medication data given its relevance and complexity. First, some basic definitions.

**Orders** are outpatient and inpatient medication prescriptions entered into the electronic health record (EHR) by providers. Order data can be extracted for research from a relational database supporting the EHR. Because each electronic health system is unique, and is primarily intended for internal use, there can be wide variation in the structure and content of order data across systems. Moreover, health systems that infrequently use order data for research might have less local infrastructure or less familiarity with using them in this way.

**Dispensings** are medication prescriptions that have been filled and sold to a patient from a pharmacy. If an order represents a treatment recommendation, then a dispensing represents a next step towards following that recommendation. Data sources include internal pharmacies within integrated systems like Kaiser Permanente or within single-payer systems like Veterans Affairs (VA). Other sources include administrative insurance claims and data packages incorporating community pharmacy data (e.g., Sure Scripts®). Dispensing data are used for billing purposes and thus tend to have similar data elements and structure across source systems. This continuity lends itself to multi-site research though data availability can differ by source. For example, inpatient dispensings might be available in health systems that operate their own hospitals but will not be available in insurance claims because inpatient medications are bundled with services for payment. Similarly, medications paid for out of pocket will be missing from insurance claims data but likely present in community pharmacy data packages and single-payer systems like the VA.

Both orders and dispensings contain information on the type of medication being ordered or dispensed. Some common medication identifiers include:

- **National Drug Code** (NDC) is a universal product identifier for all prescription and over-the-counter (OTC) medications in the United States. The Food and Drug Administration (FDA) maintains a directory of submitted NDCs that is publicly available and updated daily. Each NDC is specific to a manufacturer, strength, dosage form, formulation, package size and package type. While the codes provide detail about a product, they lack any sort of classification schema that could facilitate grouping. This can make the building of a comprehensive code list cumbersome as it often necessitates some degree of manual review. Moreover, the NDC is typically assigned for billing purposes when a medication is dispensed, and so it is available in dispensings and insurance claims data but not in orders data.
- **Generic Product Identifier** (GPI) is a proprietary coding schema that facilitates grouping. GPI hierarchically organizes drugs based on seven increasingly detailed levels of information starting with primary therapeutic use. This system makes it easy to identify a group or class of related products, such as vaccines or toxoids, but is only available with a license.
- **Generic medication name** is typically present in both order and dispensing data. It can be populated in a variety of ways, e.g., as a free-text field entered by a provider or selected from a drop-down menu. Pulling medication data based on generic name requires a priori knowledge of all possible medication names in circulation during the period of interest. It also necessitates accounting for different spellings and formats (e.g., for combination medications) when executing the data pull to ensure the widest net is cast. Medications identified based on generic name should be manually reviewed by a pharmacy expert after extraction, which can be time-intensive.

Each site’s experience using medication data for research will likely vary. This is a good thing, as it forces all sites to revisit common assumptions about their data that are often taken for granted. It also presents an opportunity for collaboration, as sites with more experience can mentor those with less experience. All sites, in turn, are bound to learn something new and valuable along the way and feel more engaged in the work.

**Step 2: Get to know your data**

Once you know the landscape, it’s time to drill into the details. We did this by compiling a list of questions based on the data matrix from Step 1 to facilitate deeper understanding of how each element is defined at each site. The goal here is to start with the most basic questions even if (especially if) the answers seem obvious or universal. This creates a common starting point irrespective of experience. Below is a list of questions that sites can use to investigate their local medication data and guide cross-site discussion. Some questions apply to both order and dispensing data while others are specific to one type. This investigatory work is integral to identifying commonalities and differences in data across sites.

*Table 2.* Questions to ask about your data

| **Domain** | **Order Data** | **Dispensing Data** |
| --- | --- | --- |
| **Dates** | What date fields are available in your local medication data and what does each field represent? | |
| **Start Date** | Is there a date the order was placed and a date the order starts? How do they differ? Which is the preferred date for defining medication initiation? | How is dispensing date defined? Is it the day a pharmacist fills the order (e.g., puts pills in a bottle) or the day it is sold to a patient? These dates can sometimes be hours or even days apart. |
| **End Date** | Is there an order end date? How is it populated? E.g., in some systems, order end date can be retroactively populated or reset based on new provider activity (like placing a new order). | How is run-out date (the date of last available dose) commonly defined in your system? Is it in your data already or does it need to be derived? |
| **Setting** | Does your data source include both inpatient and outpatient medications, or just one? | |
| **Status** | What happens when the status of an order changes in your system (e.g., when an order moves from pending to sent or canceled)? Is a single field updated, is a new record created, something else? Are you likely to need to take steps to remove duplicates created as the status changes? | Do you have access to incomplete or unfilled prescriptions or only completed dispensings? What are the possible dispositions for dispensings in your system? |
| **Medication Identifier** | What data element(s) are commonly used in your system to identify medications? E.g., generic medication name, NDC code, other? Does your site have a license to use any proprietary databases that include medication classification schema like GPI code? | |
| **Infrastructure** | What are common issues in your medication data (e.g., missingness or incomplete capture, values changing over time, unstructured data, duplicate data)? And how are those issues commonly handled?  How “ready for research” are your medication data? What is needed to prepare them for research? | |

**Step 3: Choose your data**

With the learnings from the first two steps in mind, sites need to decide (a) which type of medication data to use, and (b) which medication identifier(s) to use. Identifier(s) will depend in part on the type of data used as not every identifier is available in both.

We recommend keeping the following domains in mind when deciding which data to use:

- **Availability.** What data are available at most, if not all, sites? For example, sites with both order and dispensing data might choose orders if that’s what is available at all sites, even if there is more local infrastructure and expertise for using dispensing data.
- **Infrastructure.** How “ready for research” is each data type at each site? Data that are systematically cleaned, routinely and frequently updated, and structured according to a set of predefined specifications are often desirable for research. Data sources lacking these qualities may still be worth pursuing if a study has the time and resources to do the additional work necessary to prepare the data for research use.
- **Setting**. Based on your research question, what type of medications are needed—inpatient, outpatient, both? Orders are a reliable source for inpatient medications while some sources of dispensing data—like insurance claims—are unlikely to capture any inpatient medications.
- **Capture.** What data, and whose data, tend to be missing from each source? For example, order data coming from an EHR will not include data from care received outside of the health system. Similarly, dispensing data coming from insurance claims will not include medications paid for out of pocket. Data capture can also vary by insurance type. Within integrated systems like Kaiser Permanente (KP) that provide both insurance and health care, we typically assume capture of data will be nearly complete, but some members can have KP insurance without using KP providers. Patients like this will not have data in the EHR and thus will not have order data.

In our study, sites with both types of medication data chose to use dispensings for the following reasons:

- **Availability**. No single type of medication data was available at all sites so sites with options could choose their preferred type based on other factors.
- **Infrastructure**. A key element needed in our study was duration of medication use. This information was readily available in dispensing data (as days’ supply) whereas it needed to be derived in order data and could be inaccurate (because people do not always fill prescribed medications or utilize all allowed refills). Moreover, dispensing data were commonly used for research at these sites and were already part of an existing common data model (CDM).
- **Setting**. Our study focused on outpatient prescribing of Benzodiazepine and Z-drug medications. As such, it was inconsequential if some sites were missing inpatient dispensings (e.g., because they were using insurance claims data) as they weren’t needed.
- **Capture**. Because order data come from the EHR, they were only available for a subset of the patient populations at sites with both order and dispensing data. Using dispensing data gave these sites access to a larger portion of the target population that wasn’t dependent on insurance type or where care was received.

As reflected in Table 1, some sites will only have one type of medication data. These sites, along with the rest, will still need to decide which medication identifier(s) to use when pulling data and constructing variables. Certain identifiers, like NDC, are only (or almost only) found in dispensing data because they are used for billing. Order data typically contain generic medication name along with internal, often proprietary, schema for classifying medications, e.g., by therapeutic class. Generic name values are relatively consistent across different EHR systems but classification schema are likely to vary. Similarly, many sources of dispensing data also include proprietary classification schema like GPI code that require a license to use. Sites will likely differ in which, if any, proprietary schema are available in their medication data. This presents challenges for data extraction and manipulation that are addressed in more detail in Step 4.

NDC can be mapped to generic medication name using the publicly available NDC directory maintained by the FDA. This lends itself to harmonization with order data where only generic medication name is available. In our study, sites using dispensings used both NDC and generic medication name to identify medications in a process described in Step 4 below. Sites using orders used generic medication name only.

It's worth noting that some EHR systems might contain a field for NDC in order data. We advise against using this field unless there is strong evidence that it is reliably populated for all types of orders. For example, in some EHR systems, NDC might only be populated for inpatient orders. In other EHR systems, NDC might be populated with a default code based on generic medication name (or internal classification code) because a single medication can map to many NDC codes. An implication of this is that the default code might not represent what gets dispensed. This may be acceptable for pulling a class of medications but would not be suitable for identifying specific drugs within a class nor for characterizing which medications are most commonly prescribed within a class.

**Step 4: Pull your data**

Now that each site has decided which data to use, it’s time to pull the data. There are two primary components to consider when pulling medication data regardless of the type: date ranges and medication identifiers.

An initial pull should cast the widest net possible both in terms of dates and the medications included. With dates, this means including medications within the observation window as well as those straddling the start or end of the observation window. It also means accounting for medications that start before the observation window but are missing an end date. We recommend including medications that start up to a year prior to the start of an observation window and then looking at the data to further refine the pull.

Casting a wide net with medication identifier means starting with the broadest list of codes or search terms and refining it based on the data extract. It is not uncommon for an initial pull to yield medications beyond the ones of interest that are subsequently dropped after review.

The exact order of operations for pulling data will vary across sites depending on factors like local database structure and capacity, data streams, and anticipated size of the study cohort. Each site should have the flexibility to apply the following operational domains in whatever way makes most sense at their site.

*Table 3.* Operational domains for pulling data

| **Domain** | **Operation** |
| --- | --- |
| **Dates** | Pull all medications that start before the end of the observation window and that end after the start of the observation window. In systems where end date can be missing, include medications that start prior to the end of the observation window. This can potentially return records that start many years prior to an IRB-approved observation window so additional parameters may be needed to stay in compliance at this step.  *Recommended quality checks:*   - Distribution of medication start date by Month/Year - Distribution of medication end date by Month/Year - Missingness across all date fields |
| **Medication Identifier** | *Using Generic Medication Name*   1. Compile a list of search terms based on generic names for medications of interest (see Step 2a in Example below for list of search terms used in our study) 2. Pull local medication data by doing a string search on generic medication name using the list of search terms 3. Review the unique combinations of generic medication name, route of administration (e.g., oral), dosage form (e.g., tablet), strength and quantity to determine what to keep versus drop   *Using NDC Code*   1. Compile a list of NDCs for medications of interest (we recommend starting with a code list from a previous study) 2. Use this NDC list to pull local medication data at each site 3. Pull additional medication data that were not captured by the NDC list by doing a string search on generic medication name using a list of search terms 4. Send any newly-identified NDCs from Step 3 along with generic name, strength, quantity, route of administration and dosage form to a pharmacy expert for review 5. Any NDCs deemed relevant in Step 4 are added to the original NDC list and redistributed to sites 6. Sites re-pull local pharmacy data based on the updated NDC list from Step 5   *Recommended quality checks*:   - Distribution of generic medication name overall and by Month/Year of medication start date - Missingness across all relevant data elements - Summary statistics (min, max, interquartile range (IQR), mean/standard deviation) for quantity (i.e., the number of medication units prescribed in the order) |

**Example: Pull all orders for Benzodiazepine and Z-drug medications in 2018 (baseline year)**

1. Pull all orders that start on or before 12/31/2018 and/or end on or after 1/1/2018
   a. Generate frequency distribution of year of order start date
      i. DROP records with order start date prior to 1/1/2017
   b. Generate frequency distribution of year of order end date
      i. DROP records with order end date after 12/31/2019
2. Perform string search on generic medication name based on list of search terms compiled by pharmacy expert
   a. Search terms: ALPRAZOLAM, CHLORDIAZEPOXIDE, CLOBAZAM, CLONAZEPAM, CLORAZEPATE, DIAZEPAM, ESTAZOLAM, ESZOPICLONE, FLURAZEPAM, LORAZEPAM, MIDAZOLAM, OXAZEPAM, QUAZEPAM, TEMAZEPAM, TRIAZOLAM, ZALEPLON, ZOLPIDEM
      i. Include spelling variations and segments of words; e.g., “LORAZ” when searching for “LORAZEPAM”
      ii. Make sure to match the case (e.g., upper or lower) of search terms with case in character field
   b. Generate frequency distribution of generic medication name overall and by month and year of order start date—is anything missing? Does anything need to be dropped?
      i. DROP records that are not medications of interest
3. RETAIN orders classified as e-prescription or normal prescription
4. RETAIN orders written in an outpatient setting by a provider
   a. DROP orders classified as patient-reported
5. RETAIN orders that have been completed, sent to the pharmacy, or dispensed (if known)
6. DROP orders with >365 or <1 days between order start and end date

**Step 5: Harmonize your data**

At this point, each site will have raw medication data that reflect the nuances of their local source systems (e.g., with unique variable names, different data types for similar elements, etc). For example, a variable for medication strength might be numeric at one site, character at another, and embedded within a text string in another. There are two potential next steps from here.

One option is to define study constructs at a high enough level for each site to program independently against their unique raw data. In this decentralized approach, collaboration is largely concentrated at the beginning (e.g., defining constructs) and end (e.g., pooling site data for analysis) with minimal oversight in between. This gives sites flexibility and control over the programming process, which can be both efficient and empowering for the individual sites. At the same time, each site is independently interpreting a potentially complex set of definitions while tailoring those definitions to their unique local data. This can make it difficult to explore observed differences in the final covariates and outcomes across sites (e.g., whether differences are due to differences in interpretations of the constructs, programming error, or true differences in the cohorts). It also means that key constructs must be well defined at the outset, with very little room for modification once final analytic data have been delivered.

Another option is to establish a set of common data elements that serve as an intermediary dataset between the raw local data and the final analytic data. In this centralized approach, collaboration is infused throughout the programming process. Sites work together to both define key constructs and identify the minimum data necessary to derive those constructs. The minimum data necessary ultimately become the common data elements comprising the intermediary dataset. The intermediary dataset facilitates cross-site QA and distributed programming (meaning the same code can be run at every site to create final covariates and outcomes because the underlying data are now the same). It also creates an “audit trail” between local data and the final analytic data that is useful for exploring observed differences across sites. Having a common intermediary dataset also gives analysts flexibility to explore different ways to operationalize key constructs even after final analytic data have been delivered.

We recommend the centralized approach of transforming local medication data into a set of common data elements like the ones in Table 4. These common data elements represent the minimum data necessary to create the covariates and outcomes needed for analysis. This list is meant to be a starting point and isn’t exhaustive. Table 4 outlines how each common data element can be created from either dispensing or order data.

*Table 4.* Medication Common Data Elements

| **Data Element** | **Variable Name** | **Dispensing Data** | **Order Data** | **Data Type** |
| --- | --- | --- | --- | --- |
| Start date of medication | rxdate | date the fill was dispensed (i.e., either pills were put in a bottle or the dispensing was received by patient) | date the medication order starts | Date |
| End date of medication | rxenddate | *See Step 5.a. below* | date the medication order is to end or did end (if retroactively populated/updated) | Date |
| Generic name of medication | genericname | generic name of medication | generic name of medication | Character |
| Days’ supply of medication | rxsup | number of days of medication supplied | *See Step 5.b. below* | Numeric |
| Maximum refills allowed (if applicable) | refills | N/A | number of refills allowed for this prescription as entered by the prescriber | Numeric |
| Number of units of medication | rxamt | number of medication units (e.g., pills) dispensed | number of medication units (e.g., pills) to be dispensed as entered by the prescriber | Numeric |

Several of the data elements in Table 4 warrant a bit more context and explanation.

*Step 5.a.* Calculate medication end date.

*Context*: Dispensing data often include dispensing date (e.g., rxdate in Table 4) and days’ supply (e.g., rxsup) but not an end date (also known as “run-out date” or the date of last available dose in the dispensing).

*Recommendation*: We recommend calculating the date of last available dose based on dispensing date and days' supply (i.e., rxdate + rxsup -1). We subtract one day because we assume the first dose is taken the day the medication is dispensed.

*Step 5.b.* Calculate days’ supply.

*Context*: Order data often do not include days’ supply. Deriving days’ supply from order data can be challenging for the following reasons:

- Refills are subsumed within a single order (e.g., an order with 3 refills is represented as one record). This means the order end date often reflects the expected run-out date of the last available refill. As such, a single order with multiple refills can span weeks, months, or even years. It also means the duration of the initial fill and each refill within the order must be derived.
- Order end date can have high rates of missingness in some systems. It can also be retroactively populated or updated based on new provider activity (like placing a new order), making it difficult to know what the end date represents.
- Dose information (i.e., how many units of a medication to take per day) often must be mined from a free-text field called the SIG (instructions on how the medication should be used; e.g., “take 1 tablet twice a day for 30 days”).

*Recommendation*: Days’ supply can be calculated from order data in several ways, each with advantages and limitations. We took Approach A in our study due to time and resource constraints.

Approach A: count the number of days between order start date and order end date (plus one to account for use on both the start and end dates).

Advantages:

- Easy to program
- Easy to explain and understand

Limitations:

- Requires non-missing start and end dates
- Order start and end date are imprecise proxies for medication use. Orders can be placed days or even weeks before a medication is dispensed and received by a patient. Similarly, in systems where end date can be retroactively populated, end date might reflect the start of a new order rather than the last day a medication was used.

Approach B: use quantity (total units ordered; e.g., 60 tablets) and the SIG (a free-text field containing instructions on how the medication should be used; e.g., “take 1 tablet twice a day for 30 days”) to calculate the intended days’ supply of each fill and refill within an order (e.g., an order for 60 tablets with instructions to take 2 tablets per day translates to a 30 days’ supply).

Advantages:

- Greater precision

Limitations:

- Resource intensive; e.g., parsing the SIGs is programmatically intensive and requires iterative manual review to ensure accuracy. There can be a very large number of SIGs especially if your study data cover a long time period.

**Step 6: Build your measures**

Hooray! You now have what you need to build covariates, outcomes, and other analytic variables. Deprescribing studies will vary in what they measure and how but here we outline two concepts that may consistently be of interest, polypharmacy and chronic use.

**Polypharmacy***.* A key to deprescribing work is understanding the number of different medications study participants are using chronically. You likely will wish to measure this at baseline, and in many studies, at the end of follow-up as well. When seeking to identify any medication that can be or is used chronically, it is not feasible to start with a list of medication names, because the number of such medications is far too large. So you will want to find some kind of coding or grouping schema to use. At the same time, it’s rare that all sites will have the same classification schema available in their raw data to facilitate high-level grouping. Instead, we relied primarily on days’ supply and secondarily on local classification schema to identify chronic medications in our cohort. Below are the steps taken with each type of data to operationalize polypharmacy. Note that in our study, the “index date” refers to the date on which a person first qualified as having chronic use of one of the medications of interest (a benzodiazepine or Z-drug). It distinguished the baseline period from the follow-up period.

Steps for implementation with dispensing data

1. Pull all dispensings overlapping with index date (including those starting or ending on index date).
2. Drop all dispensings with <28 days’ supply.
3. Flag the medication that qualified the person for cohort entry (e.g., Benzodiazepine or Z-drug). (We did this because we did not want to include the medication of interest in our count of polypharmacy. Not all studies may want to do this.)
4. Drop all dispensings in the following domains using 2-digit GPI code (if available) or local classification scheme:
   a. Vaccines, toxoids, allergenic extracts, oxytocics, local anesthetics — parenteral, general anesthetics, antiseptics and disinfectants, antidotes, diagnostic products, chemicals, and medical devices.
5. Generate count of distinct NDC codes not including the record flagged in Step 3.

Steps for implementation with order data

1. Pull all orders overlapping with index date (including those starting or ending on index date; note that we excluded orders with >365 or <1 day between start and end date).
2. Drop all orders with <28 days between start and end date.
3. Flag the medication that qualified the person for cohort entry. (We did this because we did not want to include the medication of interest in our count of polypharmacy. Not all studies may want to do this.)
4. Drop all orders in the following domains:
   a. Vaccines, toxoids, allergenic extracts, oxytocics, local anesthetics — parenteral, general anesthetics, antiseptics and disinfectants, antidotes, diagnostic products, chemicals, and medical devices.
5. Generate count of distinct generic medication names not including the record flagged in Step 3.

After implementation, as a quality check, each site shared the ten most common medications (based on the count of orders or dispensings for that medication) qualifying as chronic at their site. See Table 5 for a rank-ordered list of the top 10 chronic medications at each site in our study. The number in each cell represents the rank order of that medication within the site where 1 represents the most common chronic medication at that site and 10 represents the tenth most common chronic medication at that site. A blank cell indicates that medication was not among the top ten most common at that site.

*Table 5*. Top 10 Chronic Medications by Site

|  | **Site** | | | | |
| --- | --- | --- | --- | --- | --- |
| **Medication** | **A** | **B** | **C** | **D** | **E** |
| Atorvastatin | 2 | 2 | 1 | 1 | 3 |
| Lisinopril | 1 | 1 | 6 | 2 | 9 |
| Amlodipine | 7 | 9 | 3 | 5 | 4 |
| Levothyroxine | 3 | 3 | 2 |  | 2 |
| Omeprazole | 4 | 5 | 4 | 3 |  |
| Losartan | 6 | 7 | 9 |  |  |
| Metoprolol |  | 6 | 7 | 4 | 7 |
| Gabapentin |  | 10 | 5 | 7 | 5 |
| Simvastatin | 5 | 4 |  |  |  |
| Oxycodone | 8 |  |  |  | 1 |

**Chronic medication use**. Our study focused on individuals meeting criteria for chronic use of Benzodiazepine and Z-drug medications. Given the differences between orders and dispensings, our study developed separate operational definitions of chronic use for each type of data that could be implemented with the common data elements established in Step 5.

|  | **Dispensing Data** | **Order Data** |
| --- | --- | --- |
| Definition | At least three dispensings with at least 45 cumulative days’ supply within a 100-day period | At least three prescriptions (counting both the initial order and allowed refills) within 100 days. This could include (a) one prescription with at least 2 refills OR (b) at least three distinct prescriptions within a 100-day period |
| Caveat(s) | This definition does not account for dispensings with long durations (e.g., more than 89 days’ supply). The frequency of dispenisngs with high days’ supply varies across sites. Given the 100-day window, a single dispensing with 90 days supply can greatly reduce the chances of meeting the criteria of at least three dispensings within 100 days. In contrast, orders with long durations can still meet the orders-based definition of chronic use if the order has at least two refills. | This definition relies on several assumptions based on clinical practice and processes at each site. One assumption is that most orders will be filled and dispensed to the patient within 14 days of the date the order was issued, or it will be cancelled and rendered inactive. Another is that an order end date updated by new provider activity (e.g., issuing a new prescription or a patient reporting they no longer use the medication) is an acceptable albeit imprecise proxy for end of use. |

**Index Date.** Everyone meeting criteria for chronic use was assigned an index date. As mentioned above in our definition of polypharmacy, index date represented the date at which a person met chronic use criteria and was used to distinguish the baseline period from follow-up. As with our definition for chronic use, index date was operationalized differently depending on the type of medication data used. This was primarily because our definition of chronic use using order data did not include a days’ supply threshold. However, this decision was also based on time and resource constraints specific to our study. Below we offer both our initial (more rigorous) definition using order data and the final (more practical) definition along with a brief acknowledgement of the trade-offs.

|  | **Dispensing Data** | **Order Data** |
| --- | --- | --- |
| Definition | Date of the third (or later) dispensing that put the cumulative days’ supply at or over 45 days | Initial definition: estimated date of second refill (after initial order and first refill) OR date of third order (whichever comes first)  Final definition: the last day of the year in which chronic use criteria were met (e.g., 12/31/2018 for those qualifying as chronic users during 2018) |
| Caveat(s) | There was some debate about whether to use the start date of the dispensing or the estimated run-out date of the dispensing that tipped a person over the chronic use threshold. Ultimately, we went with the start date of the dispensing because it had a more obvious correlate in order data (i.e., order start date). That said, there are sound scientific and clinical reasons for using the run-out date instead, as it prevents data needed for cohort entry from bleeding into the follow-up period. | Our initial definition for index date with order data would have required estimating the start date of each allowed refill within an order based on limited data. Our proposed approach was to divide the number of days between order start and end date (order duration) by the number of allowed refills plus initial fill (maximum possible dispensings) (i.e., order duration / maximum possible dispensings). This assumes that all allowed refills were dispensed between order start and end date and that all allowed refills were intended to last the same number of days. While this definition is conceptually more similar to the one applied with dispensings, we ultimately abandoned it because we did not have the time and resources to test those assumptions. Instead, everyone with order data was assigned the same index date: the last day of the year within which chronic use criteria were met. |

**Lastly, iterate and collaborate!**

As stated at the outset, data harmonization is an iterative and collaborative process. You might find yourself at Step 6 trying to operationalize analytic variables, only to discover that you are missing key data elements for variable construction. It is common to revisit preceding steps and iterate as necessary. For example, this might include adding another common data element to your common intermediary dataset in Step 5 and re-pulling local medication data in Step 3 to better capture the raw data needed for the new common data element.

The steps presented in this guide describe the mechanics of data harmonization, but the success of this work depends on collaboration and teamwork. Each site is the expert on their local data, and that local expertise is essential to building an effective roadmap for harmonization. Engaging stakeholders and data partners from the outset will improve the chances of successful collaboration at every step thereafter.

For programmers, it is just as critical to engage with investigators and research staff as it is to engage with other site programmers. In our study, site programmers attended larger team meetings as well as meeting separately as a group. The former ensured we kept the bigger picture in mind while the latter gave us dedicated time to get into the weeds, work through data challenges together, and leverage each site programmer’s unique expertise and insight.
