## Supplemental Tables for "Defining Key Deprescribing Measures from Electronic Health Data: A Multisite Data Harmonization Project"

**Supplemental Online Appendix: List of Tables and Figures**

Appendix Table 1: Descriptions of healthcare systems participating in Data Harmonization Workgroup

Appendix Table 2: Definitions of inclusion and exclusion criteria and lists of diagnosis codes and medications

Appendix Table 3: Characteristics of people age 65 or older with chronic benzodiazepine or Z-drug use at 5 US healthcare systems

Appendix Table 4a: Comparing chronic use defined from medication orders vs. dispensings for the same population

Appendix Table 4b: Comparing chronic use defined from medication orders vs. dispensings, stratified by number of medication orders

Appendix Table 5: Comparing measures of discontinuation defined from orders vs. dispensing data for the same population

Appendix Figure 1a: Illustration of fixed time point definition of discontinuation, without halo

Appendix Figure 1b: Illustration of fixed time point definition of discontinuation, with halo. In the top row, representing no discontinuation, there are dispensings or orders within the 30-day halo period around the fixed point. In the bottom line, representing discontinuation, there are no dispensings or orders within the 30-day halo period around the fixed point. Note: the 30-day period does not have to be symmetric around the fixed point; it represents any 30-day period that contains the fixed point within it.

Appendix Figure 2. Prevalence of Benzodiazepine and Z-drug Use in People Aged ≥65 Within 5 U.S. Healthcare Systems, 2018.

Appendix Figures 3a-3f: Results of Applying Inclusion and Exclusion Criteria at Each Site

Appendix Figure 4: Impact of Adjusting for Medication Stockpiling on Estimated Discontinuation Rates (Using Gap-Based Definition)

**Appendix Table 1: Descriptions of healthcare systems participating in Data Harmonization Workgroup**

| **Healthcare system** | **Location or region** | **Number of patients or members** | **Type of system** | **Data available** |
| --- | --- | --- | --- | --- |
| Kaiser Permanente Washington (KPWA) | Northwest US | ~680,000 | Integrated healthcare system | Both clinical data and claims/administrative data (including claims for care outside KP facilities). Both medication orders and dispensings. Uses an Epic electronic medical record. |
| KP Colorado (KPCO) | Colorado | ~569,000 | Integrated healthcare system | Same as KPWA. |
| University of Pennsylvania | Northeast US | ~117,000 inpatients  ~4.4 million outpatients | Academic healthcare system | Clinical data (EHR) including encounter, diagnosis, and medication data. Only medication orders. Uses an Epic electronic medical record. |
| Duke University | Southeastern US | >65,000 hospital discharges and > 4 million outpatient visits per year;  > 1 million older adult patients | Academic healthcare system | Clinical data (EHR) including encounter, diagnosis, and medication data (orders and dispensings). Uses Epic electronic medical record (modules include Clin/Doc, Ambulatory, ASAP, Willow and Medlink, providing access to inpatient, outpatient and emergency department care and pharmacy data).  Medication dispensing data available via Surescripts (derived from pharmacies and prescription benefit managers). |
| Durham Veterans Administration Healthcare System (VAHCS) | Southeastern US | ~63,000 | US government- funded system | Encounter, diagnosis, and medication data entered into VA’s EHR, Computerized Patient Record System, and stored in VA’s Corporate Data Warehouse. |

**Appendix Table 2: Definitions for inclusion and exclusion criteria and lists of diagnosis codes and medications**

| **Characteristic** | **Definition and code or medication list** |
| --- | --- |
| Active engagement with and/or enrollment in healthcare system | For Kaiser Permanente sites (integrated health care systems): continuously enrolled in group practice division (receiving care within Kaiser clinics) for the year of interest.  For other systems: at least 1 visit to a primary care clinician during the year of interest.  This criterion was applied to each year from 2017-2019. |
| Age 65 or older |  |
| Benzodiazepines and sedative hypnotics: medication list | **Benzodiazepines:**  Alprazolam  Chlordiazepoxide  Clonazepam  Clorazepate  Diazepam  Estazolam  Eszopiclone  Flurazepam  Lorazepam  Oxazepam  Quazepam  Temazepam  Triazolam  **Z-drugs:** Eszopiclone, Zaleplon, Zolpidem |
| Chronic use of benzodiazepine or other sedative-hypnotic (“Z drug”) | For sites with dispensing data: ≥3 dispensings and ≥45 days’ supply during a 100-day period. We focused on whether the person met criteria in 2018 and considered dispensings in 2017 and 2018 when assessing criteria. “Index date” is defined as the date of the final dispensing by which the criteria are met.  For sites with orders data: ≥3 medication orders (counting both the initial order and any refills it allows) in a 100-day period. We focused on whether the person met criteria in 2018 and considered orders from 2017 and 2018 when assessing criteria. “Index date” was assigned as 1/1/2019 for all individuals meeting chronic use criteria during 2018. |
| No serious mental illness at baseline | No diagnosis code for serious mental illness and no dispensing of antipsychotic medication during the baseline period, that is:   1) For sites using dispensing data, during the 1 year prior to index date, and 2) For sites using orders data, during the period from 1/1/2018-12/31/2018. |
| No significant cognitive impairment at baseline | No diagnosis code for dementia and no dispensing of dementia medication during the baseline period, that is:   1) For sites using dispensing data, during the 1 year prior to index date, and 2) For sites using orders data, during the period from 1/1/2018-12/31/2018. |
| Antipsychotics | See separate table for medication list.  Exclude people with 1 or more orders or dispensings for any of these medications in the 1 year prior to index date. |
| Dementia medications | Donepezil, Memantine, Galantamine, Rivastigmine  Exclude people with 1 or more orders or dispensings for any of these medications in the 1 year prior to index date. |
| Diagnosis codes for serious mental illness | See separate table. |
| Diagnosis codes for dementia | See separate table. |

**Diagnosis codes for dementia**

| **ICD-10 code** | **Description** |
| --- | --- |
| A81.01 | variant Creutzfeldt-Jakob disease |
| A81.09 | other Creutzfeldt-Jakob disease |
| A81.00 | Creutzfeldt-Jakob disease, unspecified |
| A81.2 | progressive multifocal leukoencephalopathy |
| F01.50 | vascular dementia without behavioral disturbance |
| F01.51 | vascular dementia with behavioral disturbance |
| F02.80 | dementia in other diseases classified elsewhere without behavioral disturbance |
| F02.81 | dementia in other diseases classified elsewhere with behavioral disturbance |
| F03.90 | unspecified dementia without behavioral disturbance |
| F03.91 | unspecified dementia with behavioral disturbance |
| F10.27 | alcohol dependence with alcohol-induced persisting dementia |
| F10.97 | alcohol use, unspecified with alcohol-induced persisting dementia |
| F13.27 | sedative, hypnotic or anxiolytic dependence with sedative, hypnotic or anxiolytic-induced persisting dementia |
| F13.97 | sedative, hypnotic or anxiolytic use, unspecified with sedative, hypnotic or anxiolytic-induced persisting dementia |
| F18.17 | inhalant abuse with inhalant-induced dementia |
| F18.27 | inhalant dependence with inhalant-induced dementia |
| F18.97 | inhalant use, unspecified with inhalant-induced persisting dementia |
| F19.17 | other psychoactive substance abuse with psychoactive substance-induced persisting dementia |
| F19.27 | other psychoactive substance dependence with psychoactive substance-induced persisting dementia |
| F19.97 | other psychoactive substance use, unspecified with psychoactive substance-induced persisting dementia |
| G10 | Huntington's disease or chorea with dementia |
| G30.0 | Alzheimer's disease with early onset |
| G30.1 | Alzheimer's disease with late onset |
| G30.8 | other Alzheimer's disease |
| G30.9 | Alzheimer's disease, unspecified |
| G31.01 | Pick's disease |
| G31.09 | other frontotemporal dementia |
| G31.1 | senile degeneration of brain < not elsewhere classified |
| G31.83 | dementia with Lewy bodies |

**Diagnosis codes for serious mental illness**

| **ICD-10 code** | **Description** |
| --- | --- |
| F20.x | Schizophrenia, schizophreniform disorder |
| F22 | Delusional disorders |
| F23 | Brief psychotic disorder |
| F24 | Shared psychotic disorder |
| F25 | Schizoaffective disorders |
| F28.x | Other psychotic disorder not due to a substance or known physiological condition |
| F29.x | Unspecified psychosis not due to a substance or known physiologic condition |
| F30.2 | Manic episode, severe with psychotic symptoms |
| F31.2 | Bipolar disorder, current episode manic severe with psychotic features |
| F31.5 | Bipolar disorder, current episode depressed, severe, with psychotic features |

**Antipsychotic medication list**

Chlorpromazine

Droperidol

Fluphenazine

Haloperidol

Loxapine

Methotrimeprazine

Molindone

Perphenazine

Pimozide

Thioridazine

Thiothixene

Trifluoperazine

Aripiprazole

Asenapine

Brexpiprazole

Cariprazine

Clozapine

Iloperidone

Lumateperone

Lurasidone

Olanzapine

Paliperidone

Pimavanserin

Quetiapine

Risperidone

Ziprasidone

**Appendix Table 3: Characteristics of people age 65 or older with chronic benzodiazepine or Z-drug use at 5 US healthcare systems^1^**

| Characteristic | KPWA^2^  N=681  % | KPCO  N=2088  % | Duke^3^  N=2122  % | Durham VA^3^  N=431  % | Penn  N=1453  % |
| --- | --- | --- | --- | --- | --- |
| Age, mean (SD), years | 73.2 (6.4) | 73.1 (6.3) | 73.1 (6.6) | 71.5 (5.2) | 73.3 (6.8) |
| Age, years |  |  |  |  |  |
| 65-74 | 65 | 66 | 66 | 79 | 64 |
| 75-84 | 28 | 27 | 26 | 17 | 28 |
| 85+ | 7 | 7 | 8 | 4 | 8 |
| Gender: Male | 33 | 40 | 34 | >97 | 32 |
| Race |  |  |  |  |  |
| White | 92 | 92 | 91 | 85 | 85 |
| African American | 3 | 2 | 8 | 15 | 11 |
| Asian | 3 | 1 | 1 | <3 | 1 |
| Ethnicity: Hispanic | 3 | 7 | <1 | <3 | 2 |
| BMI: Obese | 34 | 26 | 34 | 43 | 28 |
| Anxiety | 57 | 42 | 31 | 66 | 31 |
| Insomnia | 4 | 31 | 10 | 14 | 27 |
| Seizure disorder | 3 | 2 | 1 | <3 | 1 |
| Any of these indications | 60 | 62 | 43 | 77 | 51 |
| Depression | 47 | 33 | 20 | 37 | 17 |
| Diabetes | 18 | 19 | 16 | <3 | 17 |
| Chronic obstructive pulmonary disease | 16 | 13 | 13 | 18 | 10 |
| Current smoker^4^ | 10 | 5 | 9 | 19 | 7 |
| Chronic opioid use | 22 | 14 | 19 | 22 | 19 |
| Number of other chronic medications^5^ |  |  |  |  |  |
| 0 | 6 | 5 | 3 | 4 | 14 |
| 1-5 | 58 | 64 | 55 | 35 | 67 |
| 6-10 | 31 | 27 | 35 | 36 | 16 |
| ≥11 | 5 | 3 | 8 | 25 | 3 |
| Type of sedative-hypnotic |  |  |  |  |  |
| Only benzodiazepine | 83 | 73 | 61 | 66 | 65 |
| Only Z-drug | 9 | 19 | 20 | 20 | 24 |
| Both | 8 | 7 | 20 | 14 | 10 |
| Duration of sedative-hypnotic use in prior year |  |  |  |  |  |
| Mean (SD), days | 249 (108) | 275 (132) | 323 (63) | 331 (54) | 310 (151) |
| ≥ 270 days | 53 | 58 | 86 | 90 | 63 |

1. Results are shown as percentages unless otherwise stated. Percentages are out of nonmissing values. The proportion (%) of missing data for various characteristics is as follows for KPWA, KPCO, Duke, Durham VA, and Penn, respectively: for race, 2, 14, 1, < 3 and 1%; for ethnicity, 2, 10, <2, 0 and 1%; and for BMI, 1, 24, 5, <4 and 0%. 1 person was missing gender at KPCO. For other characteristics there were no missing data.
2. At KPWA, we created cohorts of people with chronic benzodiazepine or Z-drug use based on dispensing data and separately, orders data. This table shows characteristics of the cohort defined using dispensing data.
3. Duke and the Durham VA censored counts < 11 for this table, leading to some percentages being reported as < or > a given proportion.
4. Smoking status was defined from structured EHR data rather than diagnosis codes.
5. Count of chronic medications does not include benzodiazepines or Z-drugs.

**Appendix Table 4a: Comparing chronic use defined from medication orders vs. dispensings for the same population***

|  | Chronic benzodiazepine/Z drug use  defined from medication dispensings | | |
| --- | --- | --- | --- |
| Chronic use defined from medication orders | No | Yes | Total |
| No | 0 | 78 | 78 |
| Yes | 471 | 603 | 1074 |
| Total | 471 | 681 | 1152 |

*The population is KPWA members who met eligibility criteria and qualified as having chronic use of benzodiazepines and/or Z drugs based on either medication orders or dispensings.

**Appendix Table 4b: Comparing chronic use defined from medication orders vs. dispensings, stratified by number of medication orders***

| Number of distinct benzodiazepine/Z drug orders per patient | Total number meeting orders criteria for chronic use | Number also meeting criteria based on dispensings | Proportion meeting dispensings criteria |
| --- | --- | --- | --- |
| 1 | 458 | 169 | 37% |
| 2 | 80 | 40 | 50% |
| 3 | 536 | 377 | 70% |
| 4 | 19 | 10 | 53% |

*Here, “orders” refers to separate prescription orders written for a patient in the 100-day period in which they qualified as having chronic use; it does not count any allowed refills included in each order. The population is KPWA members who met eligibility criteria and qualified as having chronic use of benzodiazepines and/or Z drugs based on either medication orders or dispensings.

**Appendix Table 5: Comparing measures of discontinuation defined from orders vs. dispensing data for the same population^1^**

| Definition of discontinuation | Proportion discontinuing, based on orders data  (N=601)  n (%) | Proportion discontinuing,  based on dispensing data  (N=601)  n (%) |
| --- | --- | --- |
| Gap-based definitions^2^ |  |  |
| Discontinued for ≥30 days | 189 (31%) | 287 (48%) |
| Discontinued for ≥60 days | 151 (25%) | 216 (36%) |
| Discontinued for ≥90 days | 130 (22%) | 174 (29%) |
| Discontinued for ≥180 days | 104 (17%) | 126 (21%) |
| Fixed point definitions, no halo applied^3^ |  |  |
| No medication on hand on Day 180 | 127 (21%) | 245 (41%) |
| No medication on hand on Day 270 | 131 (22%) | 235 (39%) |
| No medication on hand on Day 365 | 154 (26%) | 237 (39%) |
| Fixed point definitions, with halo applied^3^ |  |  |
| No medication on hand on Day 180 | 123 (20%) | 190 (32%) |
| No medication on hand on Day 270 | 127 (21%) | 194 (32%) |

1. Population is eligible patients within KPWA who met the definition of chronic benzodiazepine or Z-drug use based on both orders and dispensings. Individuals with chronic use were identified in 2018 and followed for discontinuation during the period from 1/1/2019-12/31/2019.
2. Adjusted for medication stockpiling (early dispensing overlapping a prior dispensing).
3. “Halo” refers to a 30-day period with no dispensings/orders for the medication, within which the fixed point is required to fall in order to be counted as a medication discontinuation. No estimate could be generated for Day 365 applying a halo, because we did not extract data beyond 12/31/2019. All estimates are adjusted for stockpiling.
