## Supplemental Figures for "Defining Key Deprescribing Measures from Electronic Health Data: A Multisite Data Harmonization Project"

Appendix Figure 1a: Illustration of Fixed Time Point Definition of Discontinuation, Without Halo

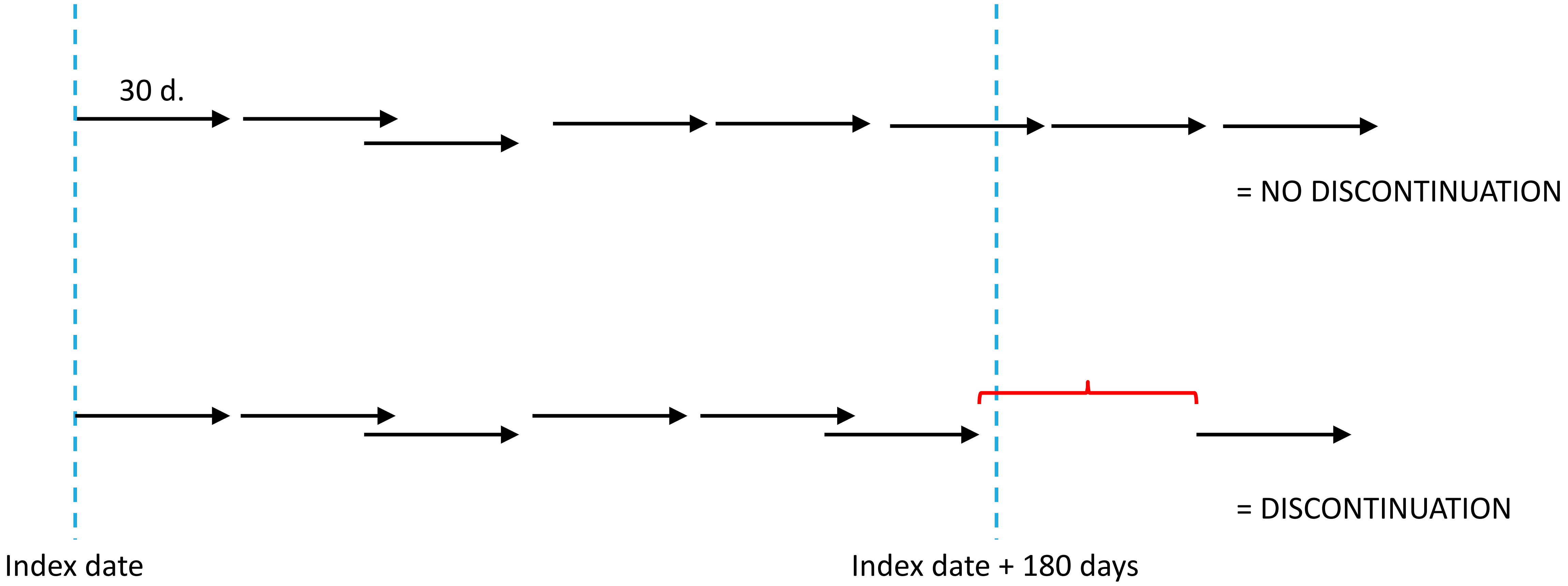

Appendix Figure 1b: Illustration of Fixed Time Point Definition of Discontinuation, With Halo

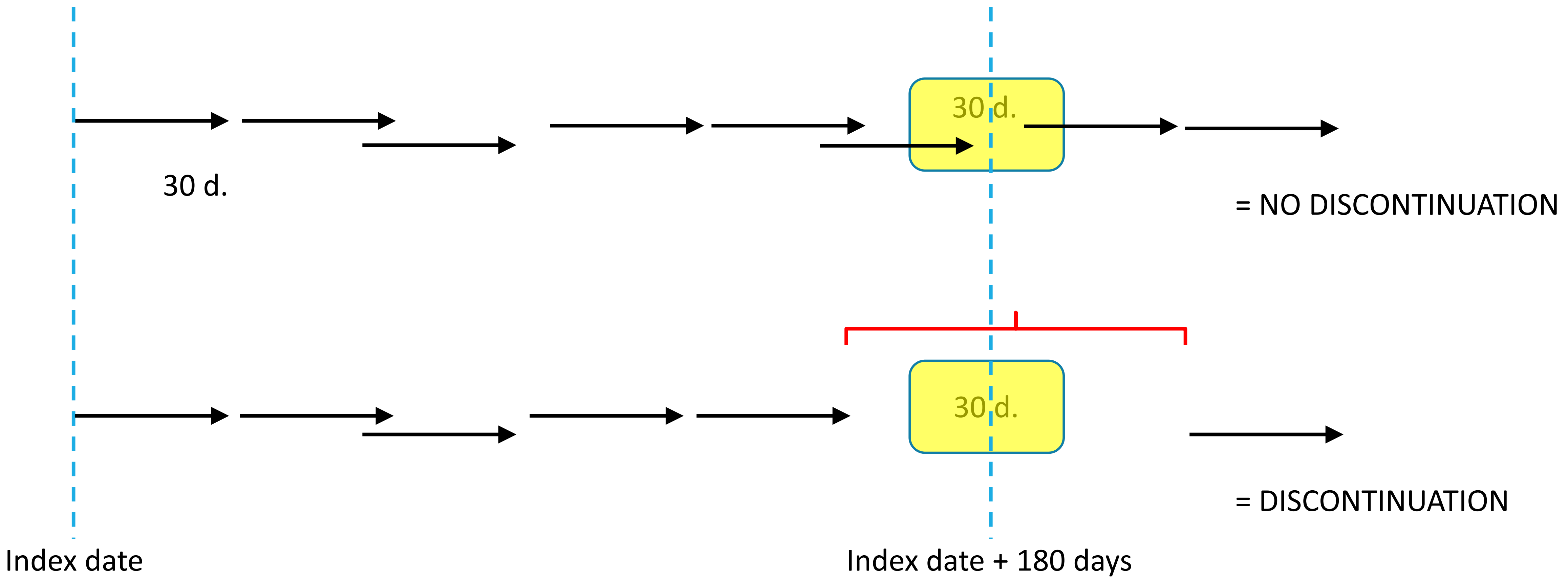


In the top row, representing no discontinuation, there are dispensings or orders within the 30-day halo period around the fixed point. In the bottom line, representing discontinuation, there are no dispensings or orders within the 30-day halo period around the fixed point. Note: the 30-day period does not have to be symmetric around the fixed point; it represents any 30-day period that contains the fixed point within it.

Appendix Figure 2: Prevalence of Benzodiazepine and Z-drug Use in People Aged ≥65 within 5 U.S. Healthcare Systems, 2018

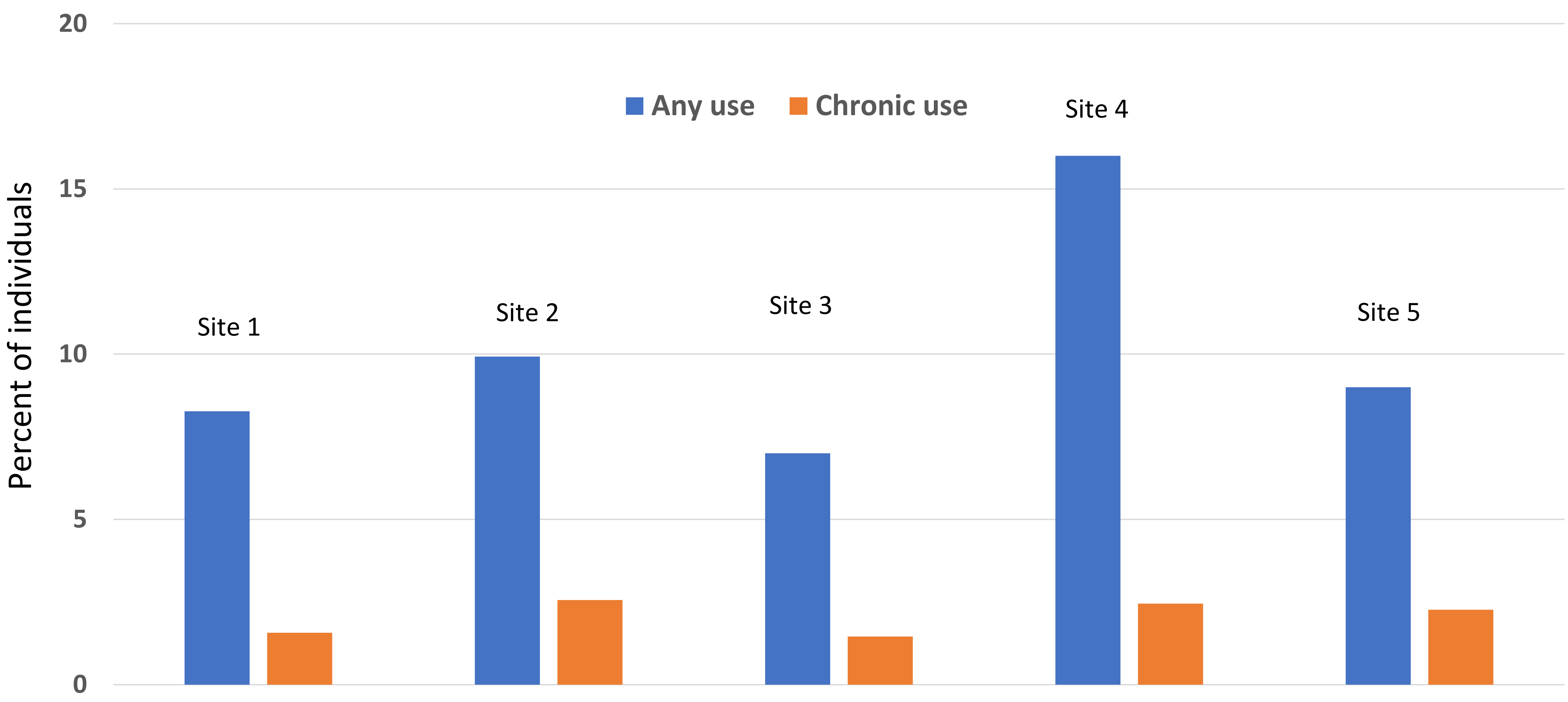

### Appendix Figure 3a: Results of Applying Inclusion and Exclusion Criteria, KPWA

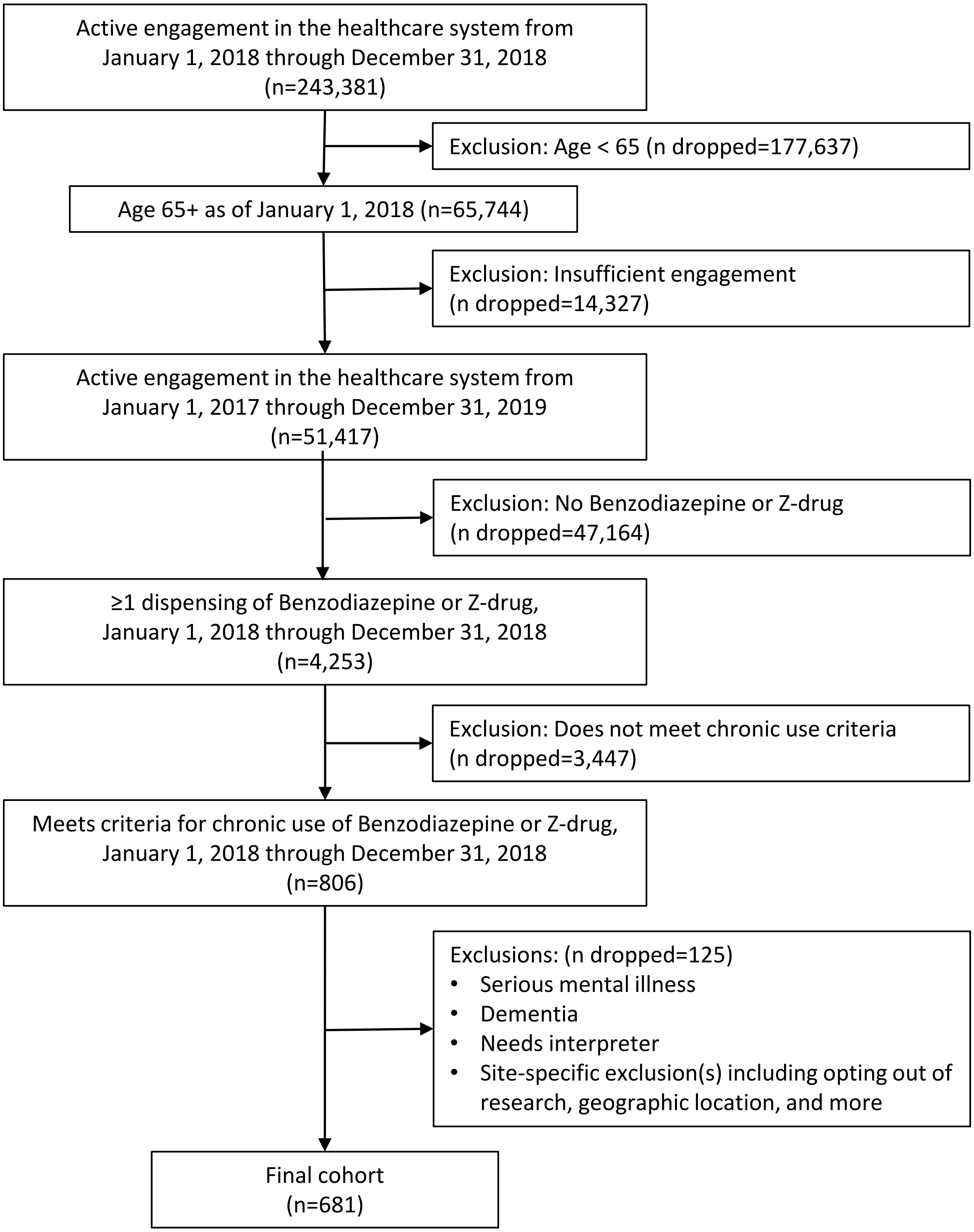

### Appendix Figure 3b: Results of Applying Inclusion and Exclusion Criteria, KPCO

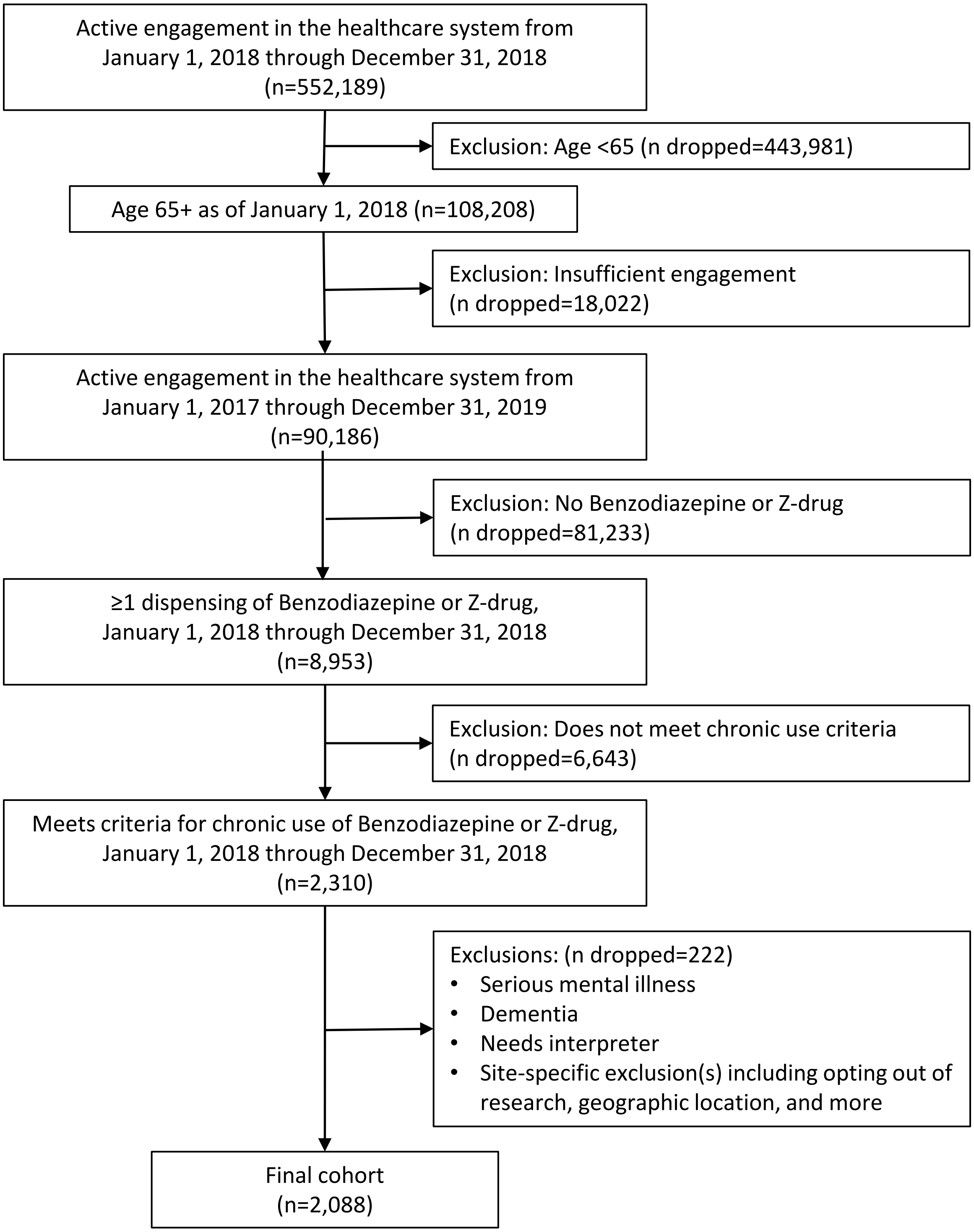

Appendix Figure 3c: Results of Applying Inclusion and Exclusion Criteria, Duke

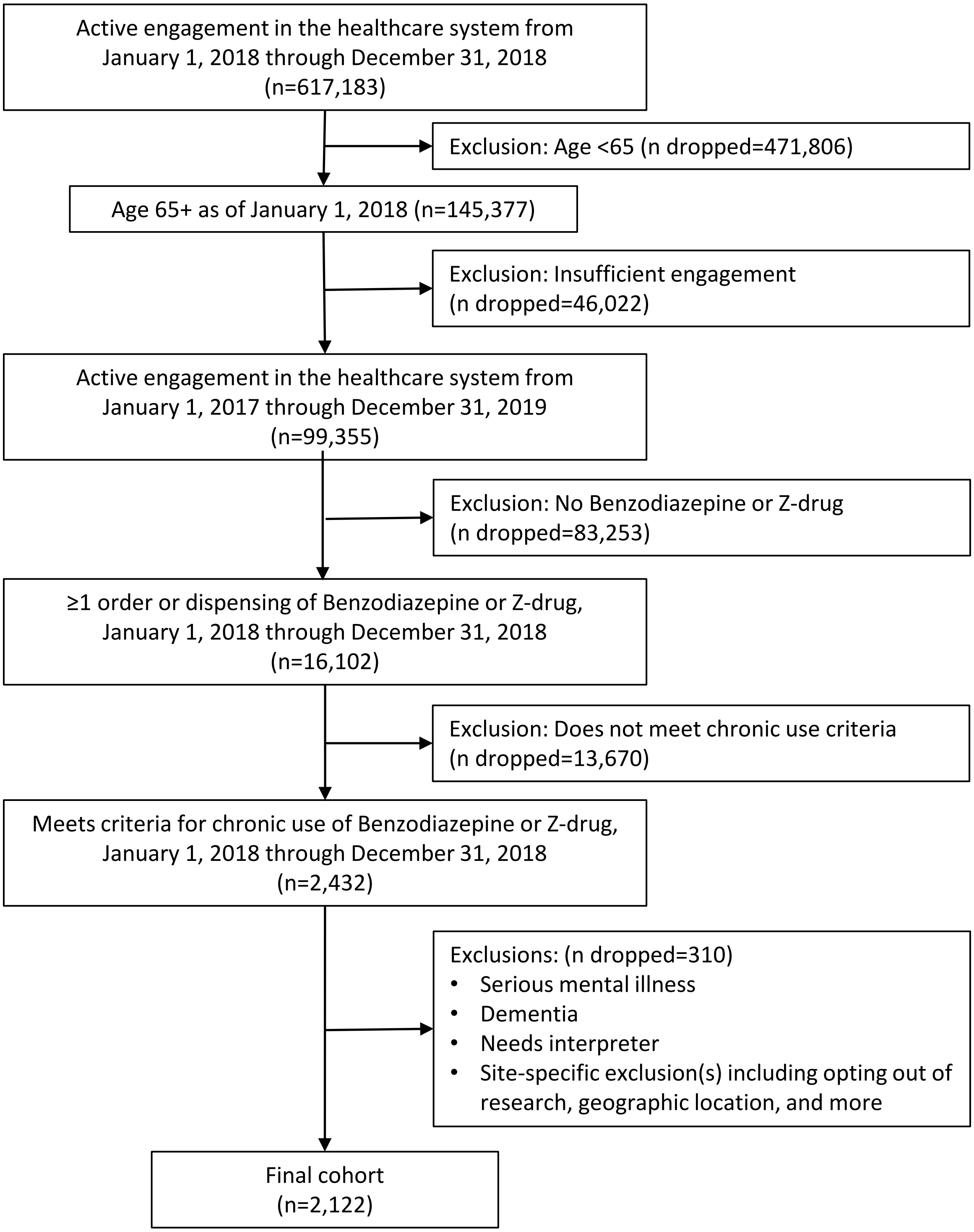

### Appendix Figure 3d: Results of Applying Inclusion and Exclusion Criteria, Durham VA

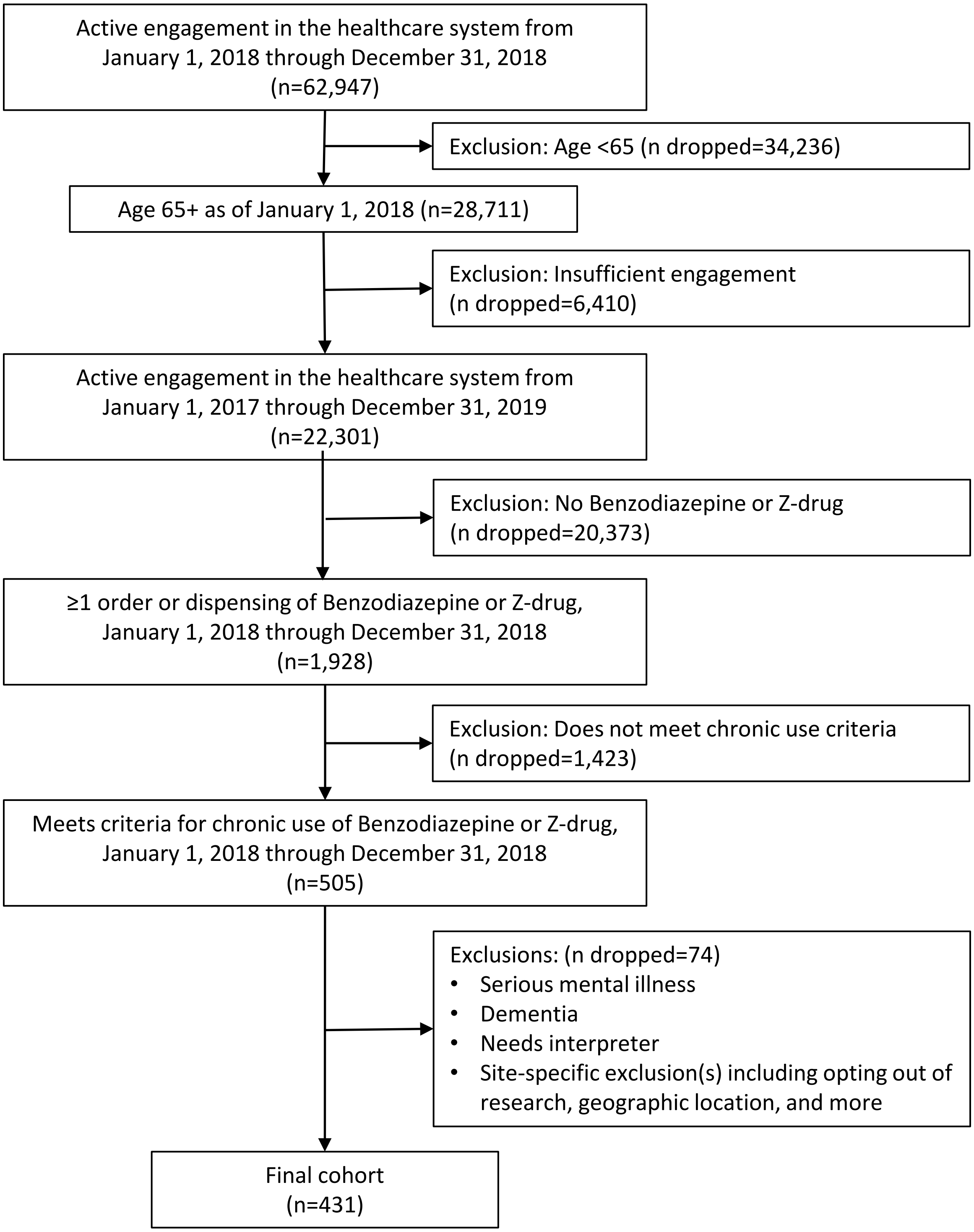

### Appendix Figure 3e: Results of Applying Inclusion and Exclusion Criteria, Penn

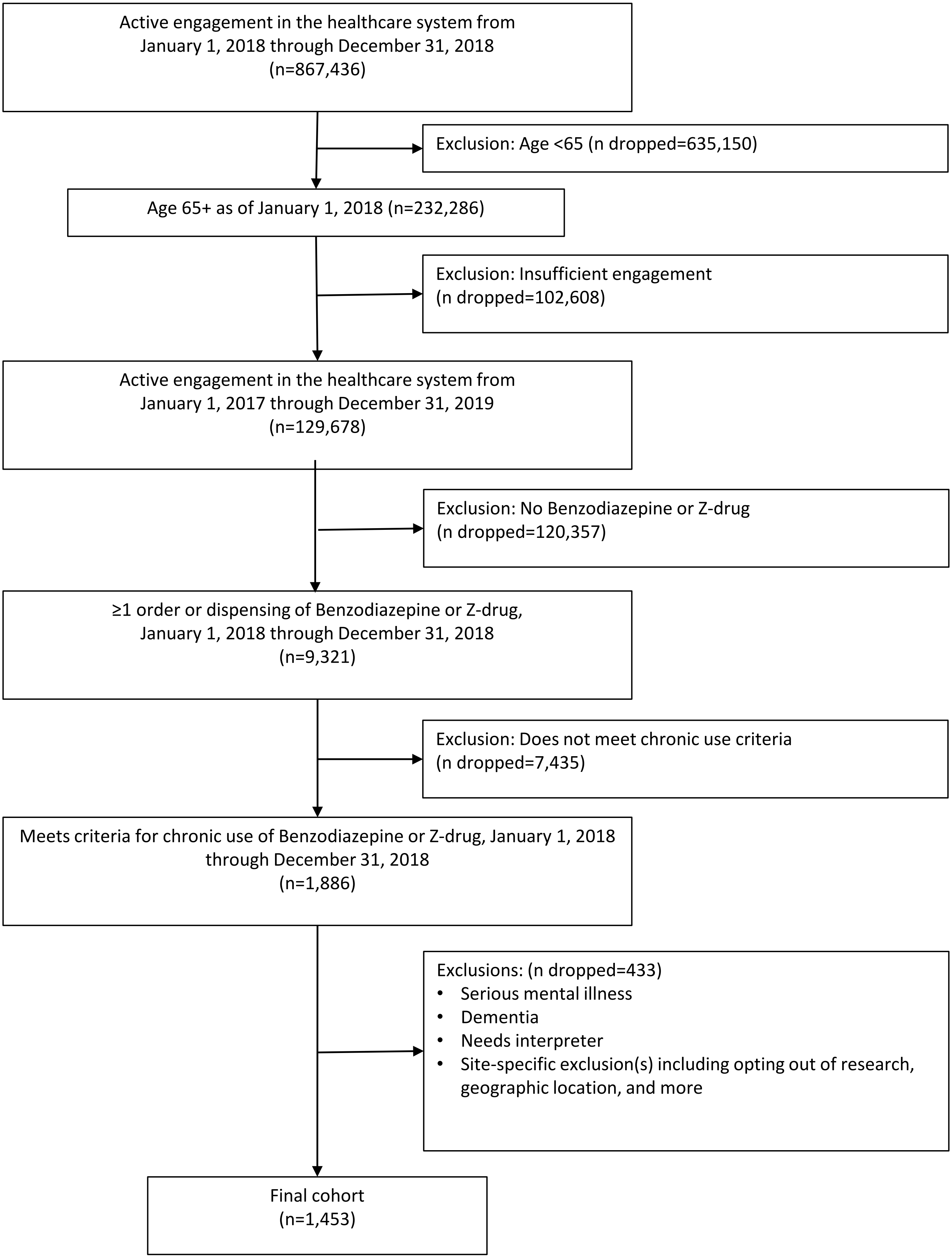

Appendix Figure 4: Impact of Adjusting for Medication Stockpiling on Estimated Discontinuation Rates (Using Gap-Based Definition)

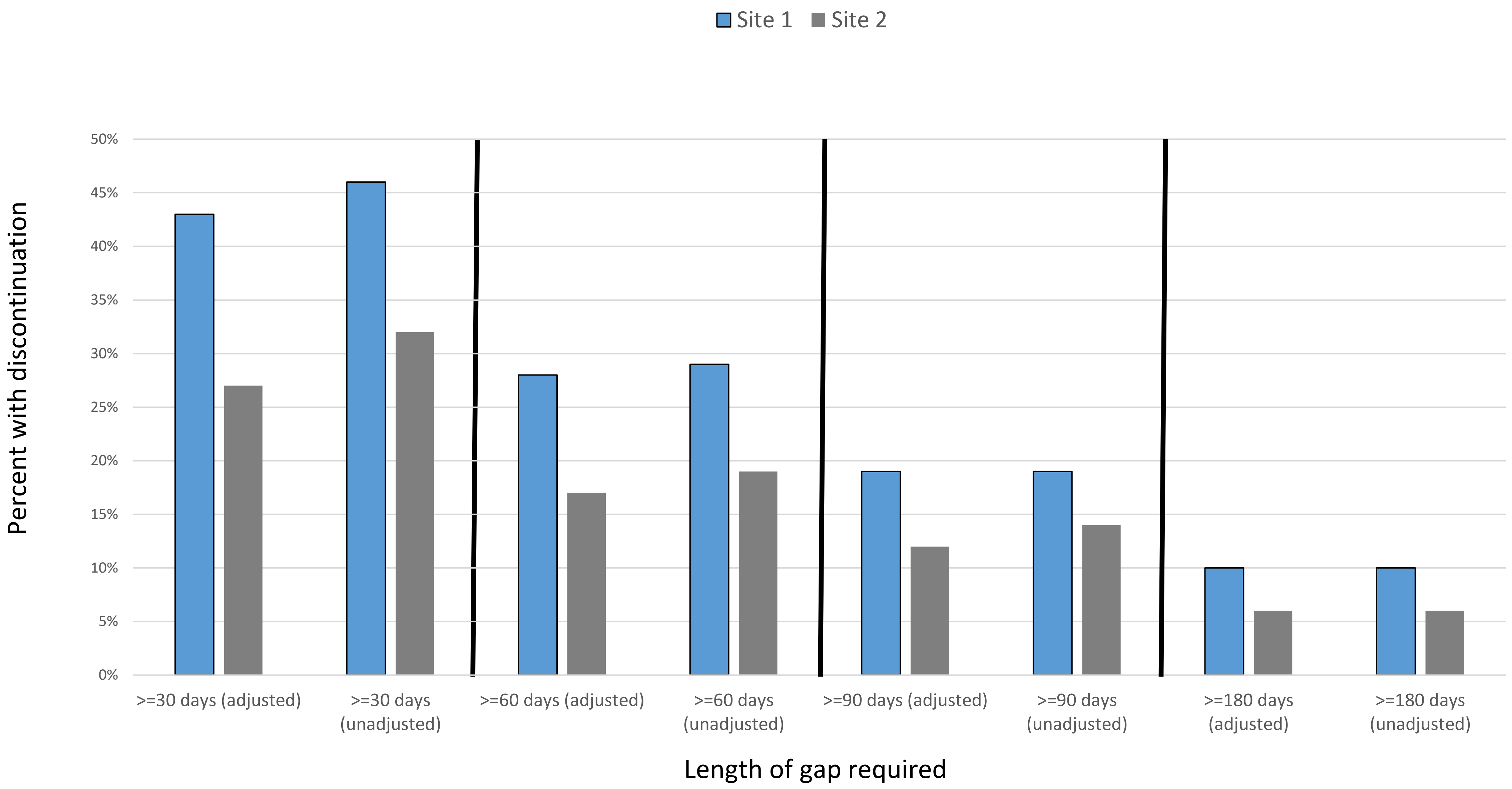
